# Strain-Level Genetic Heterogeneity and Colonization Dynamics Drive Microbiome Therapeutic Efficacy

**DOI:** 10.1101/2025.08.16.25333785

**Authors:** Kai Chen, Yina Liu, Jie Rong, Ningbin Dai, Caihua Xu, Heng Li, Ling Zhong, Baoyan Wang, Zhen Ji, Shichang Xie, Yangzuo Xu, Fulin Yang, Jing Wang, Dapeng Li, Yulan Gu, Xiumin Zhou, Yan Li, Minbin Chen, Yanan Chen, Wei Li, Zaixiang Tang, Jun Cai, Jiancheng Xu, Shuting Xia, Qimin Zhan, Zhemin Zhou

**Author notes:** These authors contributed equally (Kai Chen, Yina Liu, Jie Rong, Ningbin Dai, Caihua Xu). Correspondence: Jiancheng Xu; Shuting Xia; Qimin Zhan; Zhemin Zhou.

## Abstract

Fecal microbiota transplantation (FMT) has shown immunotherapeutic promise across multiple malignancies, yet its clinical efficacy in non-small-cell lung cancer (NSCLC) remains unexplored. Here, we report results from a single-arm trial demonstrating that FMT significantly enhances anti-PD-1 efficacy and prolongs progression-free survival in patients with advanced PD-L1-negative NSCLC. To understand the mechanistic basis of variable FMT outcomes, we developed a high-resolution strain-tracking framework and analyzed over 2,000 samples from FMT and longitudinal studies across NSCLC, melanoma, inflammatory bowel syndrome, recurrent *Clostridioides difficile* infection, type 2 diabetes, and healthy individuals.

Our analysis reveals that genetically distinct strains within the same bacterial species exert opposing therapeutic effects, explaining contradictory findings in previous reports. We discovered universal ecological principles governing strain persistence and engraftment that transcend disease contexts: engraftment success correlates with species-intrinsic fitness traits encoded in core metabolic and immune evasion pathways. Phylogenetic analysis revealed that key species segregate into functionally distinct clades with divergent clinical associations. Longitudinal tracking demonstrated that successful colonization by beneficial strain variants strongly associates with positive clinical outcomes. By integrating colonization dynamics with functional genomics, we identified 39 priority species exhibiting robust engraftment potential and strain-specific therapeutic effects as candidates for precision microbiome therapeutics. These findings establish a strain-function-efficacy paradigm that resolves inconsistent clinical outcomes in microbiome interventions and provides a framework for next-generation therapeutic development.

## INTRODUCTION

The gut microbiota exerts a profound influence on host immunity and therapeutic response, including outcomes of immune checkpoint inhibitors (ICIs) targeting the PD-1/PD-L1 axis across various cancers ^[1–4]^. Fecal microbiota transplantation (FMT) has emerged as a promising strategy to modulate the gut microbiota and enhance treatment efficacy across diverse conditions, including cancer immunotherapy^[5–7]^, inflammatory bowel syndrome (IBS)^[8–10]^, and recurrent *Clostridioides difficile* infection (rCDI)^[11–13]^. Despite its therapeutic potential, particularly in enhancing ICI responsiveness among melanoma patients^[5–7]^, FMT remains hindered by several critical limitations, including unpredictable post-transplant outcomes and the absence of standardized donor screening protocols.

Clinical studies across multiple diseases have identified correlations between specific bacterial taxa and treatment responses, yet findings remain inconsistent and sometimes contradictory across cohorts. For instance, while some studies associate *Akkermansia muciniphila* with favorable response to anti-PD-1 therapy^[1, 2]^, others reported potential side effects^[14, 15]^. Similarly, *Escherichia coli* has been implicated as both beneficial and detrimental, depending on the clinical context^[16, 17]^. These contradictory findings likely reflect functional heterogeneity that exists among genetically distinct strains within the same species—variation that is obscured by traditional species-level analyses of microbiome data^[18, 19]^.

Current strain-resolved metagenomic tools^[20, 21]^ often struggle with clinical sample complexity, particularly in detecting low-abundance organisms or differentiating closely related strains. Furthermore, existing FMT donor selection guidelines prioritize microbial diversity and pathogen exclusion^[22, 23]^, often neglecting engraftment potential—a key FMT success determinant dependent on strain-level compatibility between donor and recipient^[24–26]^.

To address these limitations, we conducted a prospective clinical trial of FMT plus immunotherapy in lung adenocarcinoma and developed a high-resolution strain-tracking framework. Applying this approach across multiple FMT cohorts and longitudinal studies spanning diverse disease settings, we demonstrate that genetic variation among conspecific strains plays a decisive role in treatment outcomes. Our findings reveal that engraftment dynamics and functional contributions of individual strains follow universal principles transcending disease-specific contexts, providing a conceptual foundation for precision microbiome therapeutics.

## RESULTS

### FMT Enhances Immunotherapy Efficacy in PD-L1-Negative Lung Adenocarcinoma

We initiated a prospective FMT plus PD-1 immunotherapy trial (CHiCTR2300076829) in patients with advanced-stage NSCLC (TNM stage III–IV) and low or absent PD-L1 expression (<1%), a population typically unresponsive to immune checkpoint inhibitors (Figure 1A). The primary cohort consisted of 13 patients who received standard first-line chemotherapy (platinum-based) combined with PD-1 blockade and oral FMT capsules from five thoroughly screened healthy donors. Three patients withdrew before completing the FMT regimen; the remaining 10 were tracked for over 11 month (355-1090 days) or until their death. One volunteer with 5% PD-L1 expression received identical therapy and was followed for 567 days, contributing microbiome samples for exploratory analysis but excluded from therapeutic efficacy assessments.

**Figure 1.**
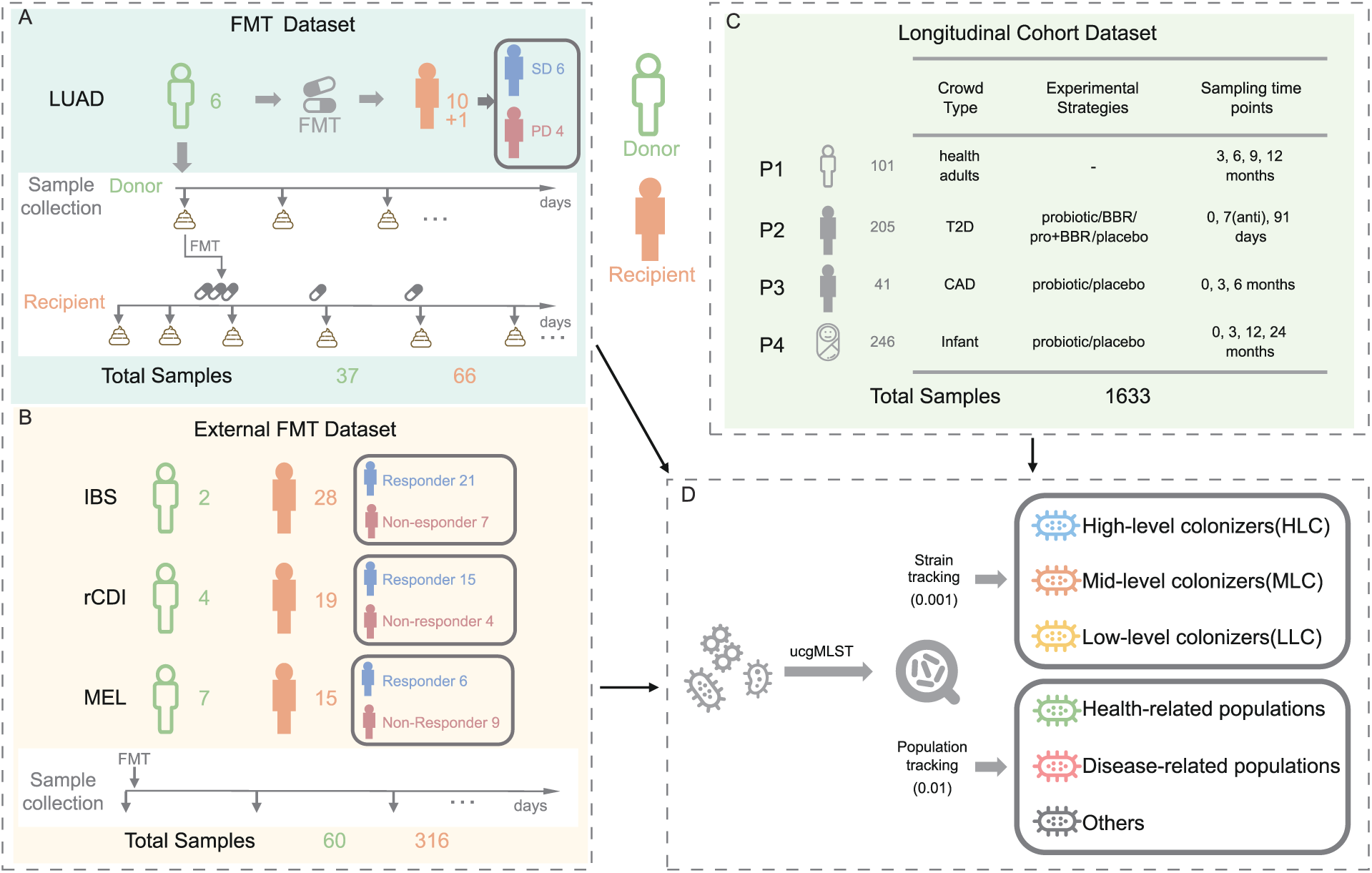
Multi-cohort study design for strain-level microbiota tracking and functional profiling across diverse clinical contexts. **A.** Primary FMT dataset from lung adenocarcinoma patients. Fecal samples from 6 donors (37 samples) and 10 recipients and 1 volunteer undergoing FMT combined with immunotherapy (74 samples) were collected longitudinally. Clinical outcomes included 6 patients with stable disease (SD) and 4 with progressive disease (PD). NSCLC, non-small cell lung cancer. **B.** External validation datasets from three published FMT studies. Irritable bowel syndrome (IBS) cohort: 2 donors, 28 recipients (21 responders, 7 non-responders); recurrent *Clostridioides difficile* infection (rCDI) cohort: 4 donors, 19 recipients (15 responders, 4 non-responders); melanoma immunotherapy (MEL) cohort: 7 donors, 15 recipients (6 responders, 9 non-responders). Total: 60 donor and 316 recipient samples across all external cohorts. **C.** Longitudinal cohort datasets for microbiome stability analysis. P1: 101 healthy adults sampled at 3, 6, 9, and 12 months; P2: 205 type 2 diabetes (T2D) patients with probiotic/berberine/placebo interventions sampled at baseline, 7 days (antibiotics), and 91 days; P3: 41 coronary artery disease (CAD) patients with probiotic/placebo treatment sampled at 0, 3, and 6 months; P4: 246 infants with probiotic/placebo treatment sampled at 0, 3, 12, and 24 months. Total: 1,633 samples. **D.** Analytical framework employing universal core gene multilocus sequence typing (ucgMLST) for high-resolution strain tracking and population-level analysis. Strain-level colonization dynamics classified transplanted microbiota into high-level, mid-level, and low-level colonizers based on persistence thresholds (0.001 nucleotide distance). Population profiling (0.01-0.02 nucleotide distance) stratified microbial populations into health-related, disease-related, and other categories.

Among ten patients completing the FMT-immunotherapy protocol, six responders achieved clinical benefit: one complete response (CR), three partial responses (PR), and two stable diseases (SD, including one patient who died of unrelated causes), yielding a 60% disease control rate (Figure 2A). In a subset of responders, tumor volumes significantly decreased and serum CEA levels declined substantially over time following treatment (Figure 2D and Supplementary Figure 1). Four non-responders experienced progressive disease (PD) and dropped out for alternative therapies.

**Figure 2.**
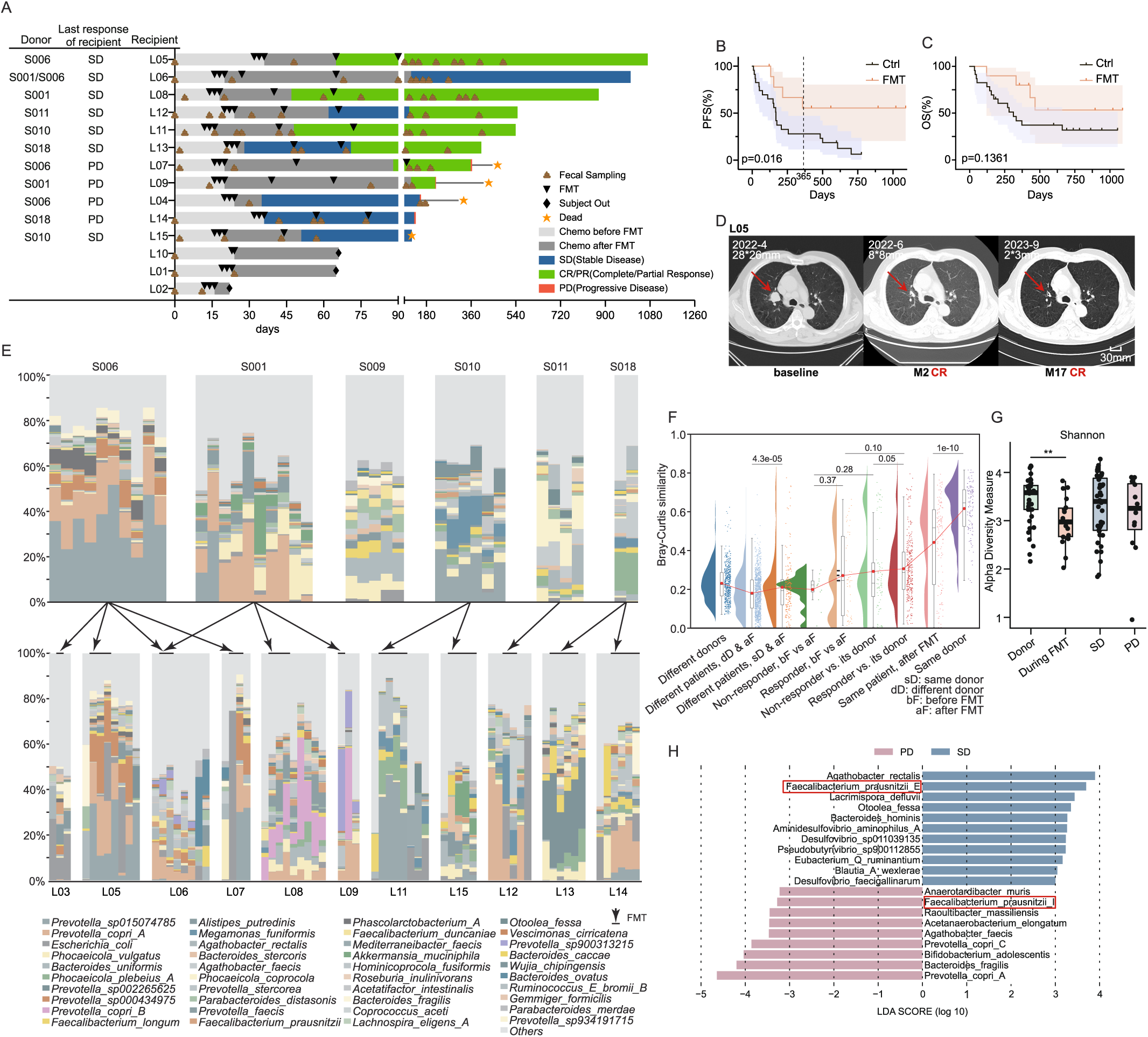
FMT treatment outcomes and microbiome dynamics in lung adenocarcinoma patients. **A.** Timeline and clinical outcomes for FMT-immunotherapy combination treatment. Individual patient trajectories showing donor assignments, FMT administration timing, chemotherapy periods, disease evaluation timepoints, and final clinical outcomes. Patients achieved either stable disease (SD, n=6) or progressive disease (PD, n=4). Symbols indicate fecal sampling, FMT treatment, dropout, death, and treatment phases. **B.** Progression-free survival (PFS) comparison between FMT-immunotherapy combination (red) and immunotherapy control (black) groups. 12-month PFS rate was extended in the FMT group (log-rank p<0.05). **C.** Overall survival (OS) comparison between FMT-immunotherapy combination (red) and immunotherapy control (black) groups. Median OS showed improvement in the FMT group (log-rank p=0.1361). **D.** Contrast-enhanced thoracic CT images of a lung cancer patient before (baseline) and after treatment (2 months and 17 months after therapy). Red arrows indicate the target lesion. **E.** Longitudinal microbiome composition at species level across donors (S006, S001, S009, S010, S011, S018) and corresponding recipients (L03, L05, L06, L07, L08, L09, L11, L15, L12, L13, L14). Stacked bar charts show relative abundances of dominant bacterial species before and after FMT treatment. **F.** Bray-Curtis dissimilarity analysis of microbiome composition. Violin plots compare beta-diversity between different sample categories, with significant differences observed between donor-recipient pairs and temporal comparisons (p-values indicated). sD, same donor; dD, different donor; bF, before FMT; aF, after FMT. **G.** Alpha diversity (Shannon index) across sample groups. Boxplots show microbiome diversity in donors, during-FMT samples, stable disease (SD), and progressive disease (PD) groups (**p<0.01). **H.** Linear discriminant analysis (LDA) effect size (LEfSe) analysis identifying differentially abundant taxa between PD (pink bars) and SD (blue bars) groups. LDA scores >2 indicate significantly enriched taxa in each clinical outcome group.

To benchmark clinical efficacy, we first compared outcomes with those from a prior clinical trial involving a matched patient cohort receiving PD-1-based immunotherapy and chemotherapy without microbiome modulation. That cohort exhibited a one-year progression-free survival (PFS) rate of 27%.^[27]^ In parallel, we retrieved data from a city-wide digital clinical data integration platform, which consolidates longitudinal electronic health records across all regional hospitals. This de-identified dataset, automatically extracted and collated over a five-year period, minimizes potential bias due to manual curation. In contrast to the historical cohorts, patients receiving FMT demonstrated markedly improved outcomes, with a 12-month PFS rate of 55% (95% CI: 23%–80%) compared to 28% (11%–47%) in the matched controls (Figure 2B, hazard ratio [HR] = 0.36, p = 0.016), reflecting a significant clinical benefit. Overall survival also trended favorably in the FMT group, although statistical significance was not reached (Figure 2C, log-rank p = 0.14). The volunteer with 5% PD-L1 expression also achieved a partial response following identical treatment. No grade ≥3 adverse events attributable to FMT were observed. All data were fully de-identified prior to access, and use of these records was approved by the relevant institutional review boards and ethics committees (ethnic: SUDA20250701H01).

### Species-Level Microbiome Analysis Reveals Limited Cross-Cohort Consistency

Longitudinal metagenomic analysis revealed stable donor microbiota compositions over time and some recipients exhibited a trend toward a microbial community composition that shifted towards the donor’s profile (Figure 2E-F). Despite established clinical efficacy in our NSCLC cohort, we observed a lack of correlation between the species-level engraftments and the treatment outcomes: there was no significant difference in the Bray–Curtis similarity between responders and non-responders before and after transplantation (Figure 2F); and all patients experienced increased alpha-diversity post-FMT, the responders have not exhibited significant greater alpha-diversities than the non-responders (Figure 2G, Supplementary figure 2). Similarly, PCoA analyses also showed that the recipients sharing similar microbiomes than the donors do not necessarily exhibit better treatment outcomes (Supplementary figure 3).

Furthermore, species-level microbiome analysis revealed concerning inconsistencies across cohorts. Linear discriminant analysis identified 26 bacterial species enriched in progressive disease patients and 39 enriched in stable disease patients in our NSCLC cohort. Notably, two closely related *Faecalibacterium* species, designated *F. prausnitzii*_E and *F. prausnitzii*_I, sharing 84.6% average nucleotide identity (Supplementary figure 4), were differentially associated with response: *F. prausnitzii*_E was enriched in stable disease/responder patients, whereas *F. prausnitzii*_I was predominantly found in progressive disease patients (Figure 2H).

To establish generalizability across disease contexts, we integrated data from three external FMT cohorts involving 62 patients: irritable bowel syndrome (IBS, n=28), recurrent *C. difficile* infection (rCDI, n=19), and melanoma (MEL, n=15), totaling 316 recipient samples (Figure 1B). We analyzed four longitudinal cohorts including 246 patients with type 2 diabetes (T2D, n=205) and coronary artery disease (CAD, n=41), plus two healthy cohorts comprising 101 adults and 246 infants (Figure 1C). The compiled dataset included 2,112 samples from 319 patients and 366 healthy controls, spanning six disease conditions.

When we extended this analysis to the three additional FMT cohorts, no single bacterial species was consistently associated with treatment response across three or more cohorts. Only 0-6 (0-5%) species were shared between any two cohorts, underscoring the limited reproducibility of species-level biomarkers (Supplementary Figure 5). Similarly, comparisons between diseased and healthy states revealed inconsistent changes in alpha diversity, indicating no universal association with health status (Supplementary Figure 6). These findings reinforce that species-level analyses fail to capture functionally relevant intra-species genomic variation, which may be the critical determinant of microbiome-mediated therapeutic outcomes.

### High-Resolution Strain Tracking Reveals Opposing Therapeutic Effects Within Species

To resolve microbial strain dynamics in complex clinical microbiomes, we developed ucgMLST, a high-resolution strain-tracking framework based on ultra-conserved single-copy genes (USCGs) (see methods, Figure 3A).

**Figure 3.**
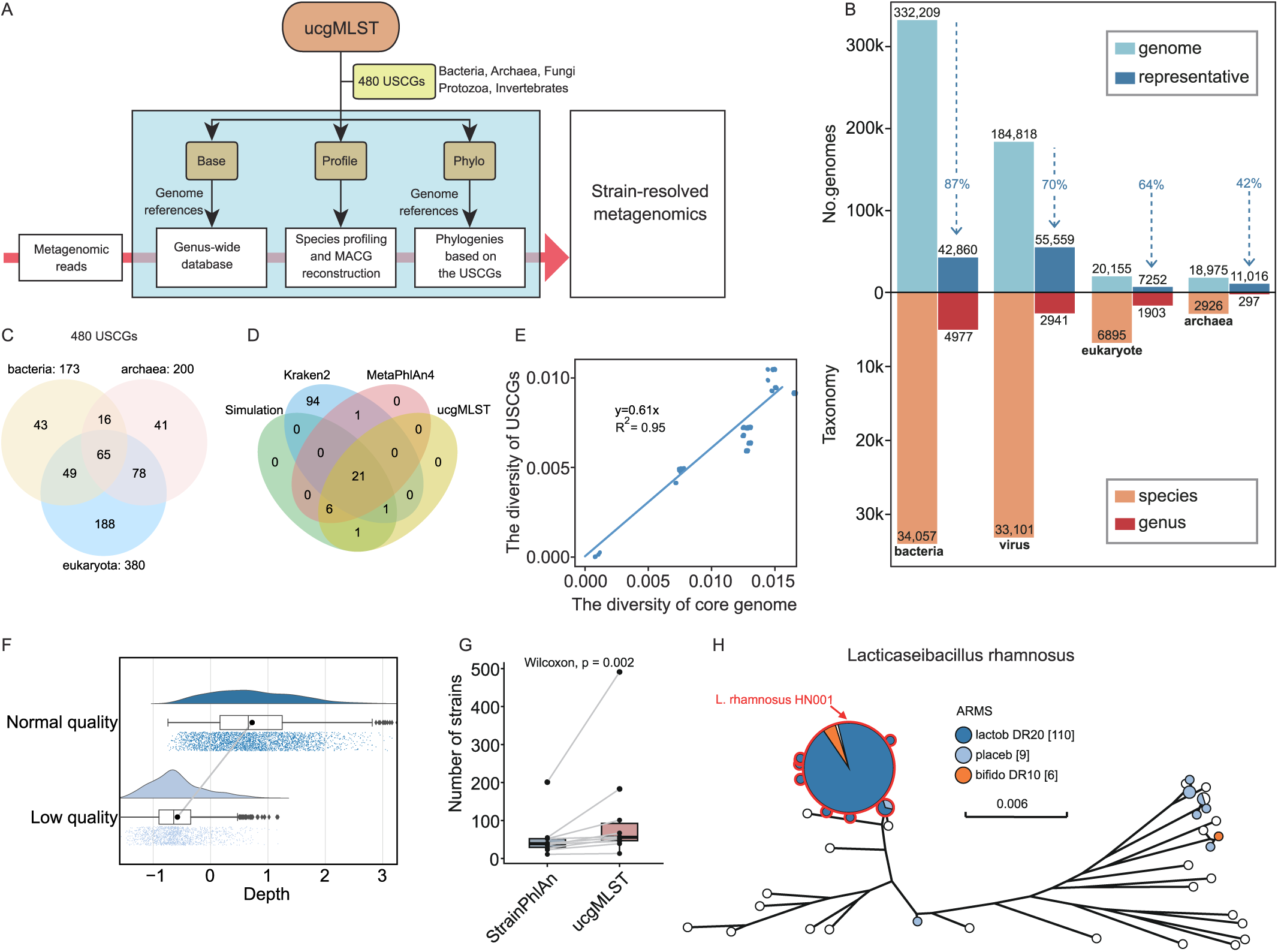
Development and validation of ucgMLST for strain-level microbiome tracking. **A.** Workflow of the universal core genome multilocus sequence typing (ucgMLST) method. The approach utilizes 480 universal single-copy core genes (USCGs) from bacteria, archaea, fungi, protozoa, and invertebrates to generate genome references, gene-wide databases, and phylogenetic profiles for strain-resolved metagenomics analysis. **B.** ucgMLST database composition showing taxonomic coverage. The database was compiled by selecting one representative for each cluster of 98% nucleotide identities from the GenBank database. The resulting representative sequences (dark blue) achieved 42-87% reduction of sizes comparing to original dataset (light blue) while maintaining the majority of genetic diversities. The species (orange) or genus (red) composition of the final reference database were shown below. **C.** Venn diagram illustrating the universal core gene set composition. The 480 universal core genes comprise 173 bacterial genes, 200 archaeal genes, and 380 eukaryotic genes, with overlapping gene families across domains. **D.** Comparison of strain identification performance between Kraken2, MetaPhlAn4, and ucgMLST using a simulated dataset with 28 *Streptococcus* species. Numbers indicate strain identification counts under normal and low-quality read coverage scenarios. **E.** Correlation analysis between the diversity of core-genome and USCGs. A strong positive correlation (R² = 0.95) supports consistency between USCGs and core genome. **F.** Violin plots show read depths of the reconstructed strains. High-quality bases were reconstructed for strains with read depths of 1.35 - 1778-folds, whereas low quality strains were reconstructed for strains with 0.1-fold depths. **G.** Comparative performance between StrainPhlAn and ucgMLST for strain detection. Box plots demonstrate significantly higher strain detection capability of ucgMLST compared to StrainPhlAn (Wilcoxon p = 0.002). **H.** Phylogenetic tree of *Lacticaseibacillus rhamnosus* strains tracked by ucgMLST. The tree shows strain relationships and identifies probiotic strain *L. rhamnosus* HN001 among T2D cohort, underscoring the detection of the same strains across individuals.

Building upon multilocus sequence typing principles, ucgMLST employs a curated set of 480 USCGs conserved across bacteria, archaea, viruses, and microbial eukaryotes (Figure 3B-C). We constructed a reference database from 556,157 high-quality genomes spanning over 76,979 species, reducing it to a compact 3.7 GB for efficient analysis. Performance benchmarking demonstrated superior capabilities compared to existing methods, with 42–87% size reduction while maintaining accuracy (Figure 3B). Each genome yielded approximately 120 kb of USCG sequence, broadly dispersed across chromosomes (Supplementary Figure S7), exhibiting sufficient sequence diversity to discriminate strains while maintaining evolutionary conservation essential for robust phylogenetic inference. Benchmark comparison with current state-of-the art methods revealed that our methodological advance enabled high species detection accuracy as well as unprecedented resolution of strain-level variation that underlies the contradictory clinical findings observed at the species level (Figure 3D-H, Supplementary Figure S8).

### Strain-Resolved Analysis Reveals FMT Colonization Dynamics

Applying ucgMLST to the lung cancer FMT cohort, we identified 48,774 unique strains across 689 microbial species, enabling quantitative assessment of strain persistence (proportion of strains shared between time points within individuals) and donor strain colonization (proportion of donor strains successfully established in recipients post-FMT, hereafter abbreviated as FMT rate). Notably, pairwise comparisons revealed excessive occurrence of closely related strains (genetic distance <0.001), particularly within samples from the same individual. This threshold, previously used in recent transmissions^[28, 29]^, was adopted to define strain-level identity in our analysis (**Figure 4A**).

**Figure 4.**
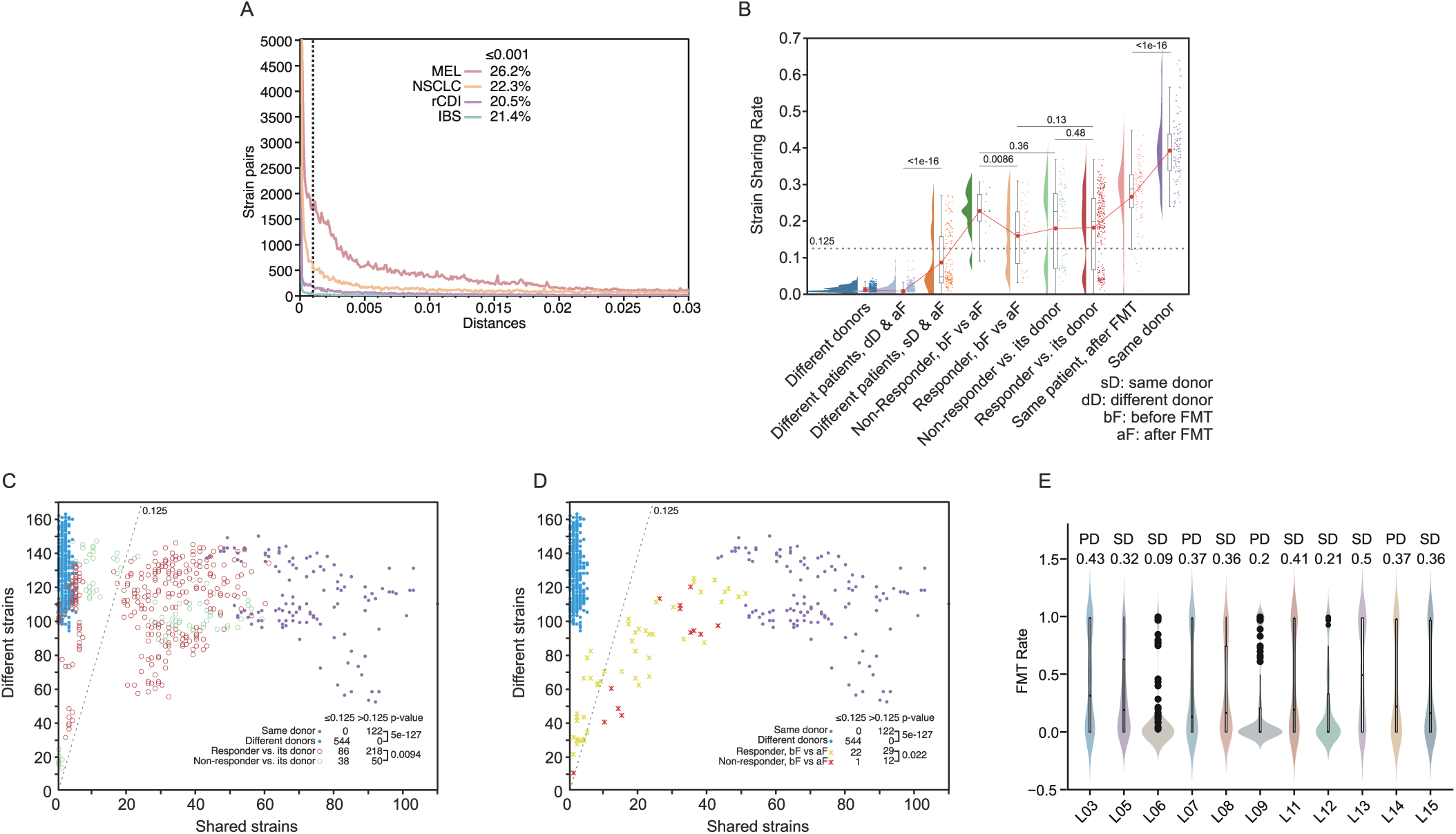
Strain-level tracking reveals FMT transmission dynamics and colonization patterns. **A.** Genetic distance distribution between strain pairs using ucgMLST. The threshold of 0.001 genetic distance (dashed line) defines strain identity, with most strain pairs showing distances below this cutoff. **B.** Violin plots showing strain sharing rates across different sample comparison categories. Violin plots show proportion of shared strain distributions for: different donors, different recipients before and after FMT, non-responder vs. responder comparisons, same patient before/after FMT, and same donor samples. Threshold of 0.125 indicates significant strain sharing (dashed line). Statistical significance indicated above comparisons. **C, D.** Strain sharing analysis between donor-recipient pairs (C) and for individual recipients (D). Scatter plot shows shared strains (x-axis) vs. different strains (y-axis) for same donor comparisons (purple), different donor comparisons (blue), responder or non-responder categories with their donors (red or green circles in C), and responder or non-responder comparing to its prior-FMT microbiome (red or yellow stars in D). **E.** Violin plots showing the FMT rates of each bacterial species for each patients by their outcome (PD, progressive disease; SD, stable disease). Bimodal distributions were observed in all recipients, with average FMT rates showing above.

Pairwise comparisons (**Figure 4B**) of the samples confirmed >1000-fold increase in strain sharing between feces from the same person, even among samples of >2 years apart, compared to those from unrelated individuals (0.23-0.63 vs <0.01 shared strains, p < 0.001). Persistence declined gradually over time: samples taken ≤16 days apart shared 0.45 strains on average, compared to 0.3 for intervals >128 days. (Supplementary Figure 9)

Notably, our comparison revealed a bi-modal distribution of strain sharing between the donor and its recipients: approximately 68% of the recipients shared 12.5% of their gut microbiome strains with the donors, whereas the remaining shared significantly lower proportions. This prompted the using of 12.5% sharing rates as a threshold for successful engraftments (**Figure 4C**.

With this threshold, donor-recipient strain sharing analysis demonstrated successful transmission events, with responders showing significantly higher successful FMTs (0.72 vs 0.57, p = 0.0094) and lower sharing with pre-treatment samples (0.57 vs 0.92, p = 0.022) compared to non-responders, suggesting the correlation between successful strain-level engraftments and the therapeutic responses (**Figure 4C and D**).

### Universal Ecological Principles Govern Strain Colonization Across Disease Contexts

Applying identical analytical pipelines to all FMT cohorts revealed variable colonization efficiencies across disease contexts, with rCDI showing the highest rate (0.24), followed by NSCLC (0.23), while IBS and melanoma exhibited lower rates (0.18 and 0.13, respectively) (Supplementary Figure 10).

However, within each cohort, strain-resolved analyses revealed a striking species-specific gradient in persistence and colonization rates that remained remarkably consistent regardless of donor identity, host condition, or disease context. For example, *Akkermansia muciniphila* strains successfully engrafted in over 80% of FMT recipients with exceptional transmission success (88% FMT rate and 100% persistence), whereas *Enterobacter kobei* exhibited poor colonization (<10%) despite repeated FMT administrations (2.6% FMT rate, 5.2% persistence) (**Figure 5A and B**).

**Figure 5.**
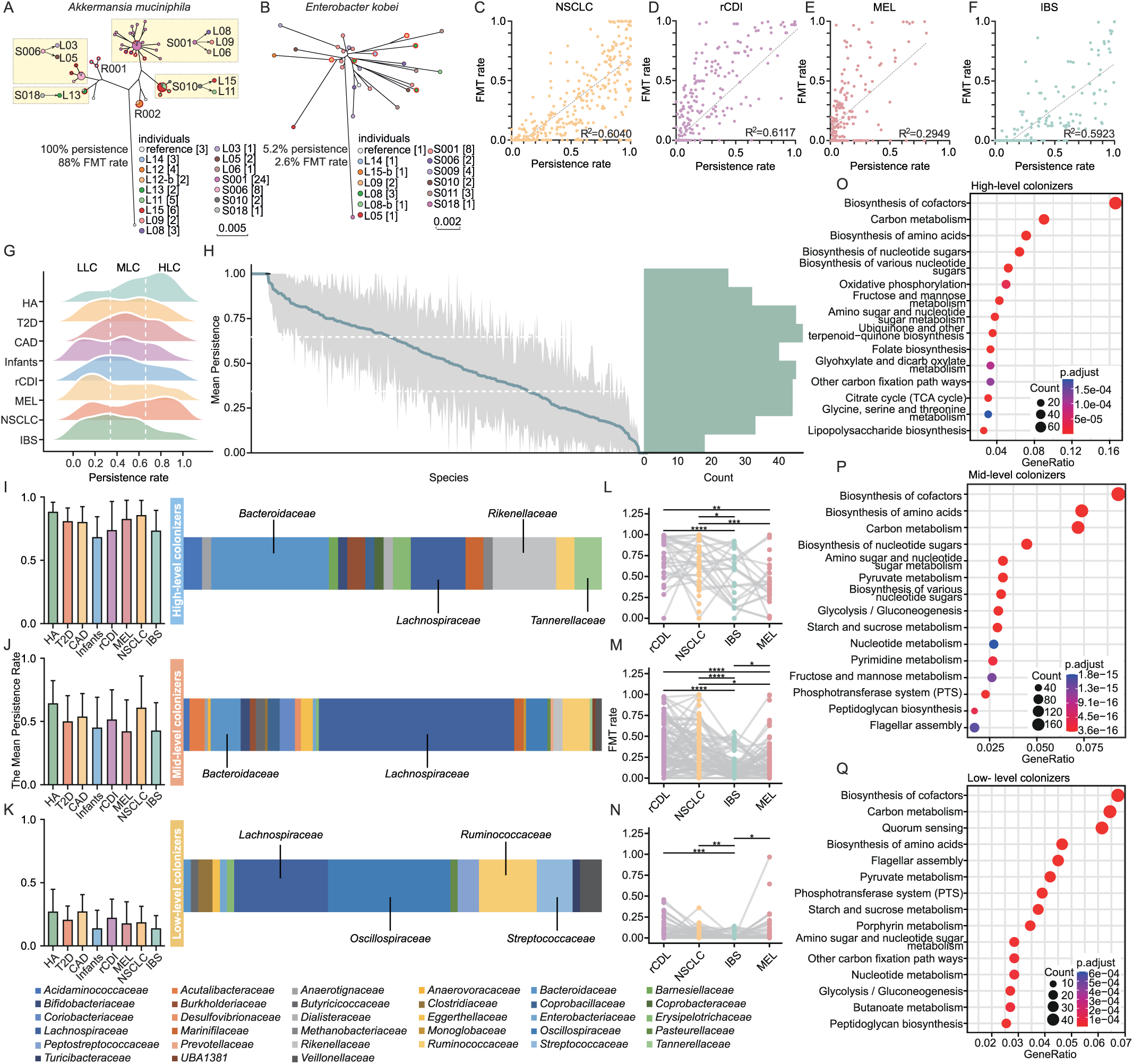
Systematic analysis of microbial colonization patterns across FMT cohorts. **A.** Phylogenetic analysis of *Akkermansia muciniphila* strain transmission. Maximum-likelihood tree based on universal single-copy core genes (USCGs) shows donor-recipient strain relationships. Yellow boxes highlight successful donor-to-recipient transmission events. Colored nodes represent different individuals (donors: S001, S006, S010, S018; recipients: L-series). Numbers in brackets indicate strain counts per individual. Scale bar = 0.005 substitutions per site. FMT transmission rate: 88%, persistence rate: 100%. **B.** Phylogenetic analysis of *Enterobacter kobei* strain transmission. USCG-based tree showing lower transmission efficiency compared to *A. muciniphila*. Colored nodes represent different individuals with transmission events indicated. Scale bar = 0.002 substitutions per site. FMT transmission rate: 2.6%, persistence rate: 5.2%. **C-F.** Correlation between FMT rates and persistence rates across four FMT cohorts. Scatter plots with linear regression lines show positive correlations for: (C) lung cancer (NSCLC) cohort (R² = 0.6040), (D) recurrent *C. difficile* infection (rCDI) cohort (R² = 0.6117), (E) melanoma (MEL) cohort (R² = 0.2949), and (F) irritable bowel syndrome (IBS) cohort (R² = 0.5923). **G.** Classification framework for bimodal persistence patterns across eight cohorts (HA, healthy adults; T2D, type 2 diabetes; CAD, coronary artery disease; Infants; rCDI; MEL; NSCLC; IBS). Ridge plots show persistence rate distributions, with vertical lines demarcating high-level colonizers (LLC), mid-level colonizers (MLC), and low-level colonizers (LLC). **H.** Species-level persistence analysis across all cohorts. Left panel shows mean persistence decay curve with 95% confidence intervals. Right panel displays histogram of species counts by persistence levels. **I-K** Taxonomic composition and persistence characteristics of colonization categories. Bar charts show mean persistence rates (left) and stacked bar charts display genus-level taxonomic composition (right) for: (I) LLC dominated by Bacteroidaceae and Lachnospiraceae, (J) MLC with mixed Bacteroidaceae and Lachnospiraceae, and (K) LLC enriched in Lachnospiraceae, Oscillospiraceae, and Ruminococcaceae. **L-N.** FMT rate comparisons across cohorts for each colonization category. Box plots show FMT rates for rCDI, NSCLC, IBS, and MEL cohorts in: (L) LLC, (M) MLC, and (N) LLC groups. Statistical significance indicated by horizontal bars and asterisks. **O-Q.** Functional pathway enrichment analysis for colonization categories. Bubble plots show KEGG pathway enrichment with gene ratios (x-axis) and statistical significance (color scale) for: (O) high-level colonizers enriched in cofactor biosynthesis and central metabolism, (P) mid-level colonizers enriched in amino acid and nucleotide metabolism, and (Q) low-level colonizers enriched in carbon metabolism and quorum sensing pathways.

Robust positive correlations emerged between species’ persistence rates and colonization likelihood across all cohorts: lung cancer (R² = 0.604), IBS (R² = 0.612), rCDI (R² = 0.295), and melanoma (R² = 0.592) (**Figure 5C-F**). This association suggests that colonization efficiency represents an emergent property of species’ high-level ecological fitness, supporting the existence of conserved ecological rules governing microbiome colonization across diseases.

### Taxonomic and Functional Stratification of Colonization Classes

To systematically analyze colonization dynamics, we stratified microbial species from all eight cohorts into three ecological classes based on persistence thresholds: high-level colonizers (HLC, persistence rate ≥ 66%), mid-level colonizers (MLC, persistence rate 33–66%), and low-level colonizers (LLC, persistence rate ≤33%) (**Figure 5G**). To eliminate cohort differences, we pooled species from all cohorts for uncertainty analysis, which indicated that the engraftment rates of specific species were not influenced by cohort variations (**Figure 5H**).

Taxonomic profiling revealed distinct compositional patterns across classes (**Figure 5I-K**). HLC were predominantly enriched in Bacteroidaceae (43.7%) and Rikenellaceae (28.1%), MLC in Lachnospiraceae (51.3%), and LLC in Oscillospiraceae (39.2%) and Ruminococcaceae (27.6%). These patterns were conserved across all cohorts, suggesting that colonization potential is encoded in microbial lineage and life-history strategy.

Temporal analysis revealed distinct lifespan dynamics across persistence classes. While overall strain persistence declined over time in all cohorts (Supplementary Figure 9), HLCs exhibited minimal decay after initial low-level drop (within 1–2 days post-FMT), maintaining stability over years, suggesting active engraftments. In contrast, LLC species showed continuous attrition, with persistence rates decreasing monotonically over time.

Cohort-specific differences in engraftment outcomes were largely explained by differential colonization of HLC and MLC taxa. The rCDI and lung cancer cohorts exhibited the highest overall engraftment rates (**Figure 5L–N**), coinciding with greatest colonization of HLC and MLC species, potentially facilitated by disrupted host microbiomes and more frequent FMT dosing.

### Functional Genomic Architecture Underlying Microbial Colonization

Functional profiling revealed distinct genomic programs underlying colonization behavior, suggesting that ecological persistence is encoded in species-specific functional repertoires. HLC species were significantly enriched for pathways associated with immune evasion and mucosal adaptation (Figure 5O), including amino-sugar metabolism, extracellular polysaccharide biosynthesis, and anaerobic respiration pathways. This signature reflects a strategy of stable niche colonization and immune modulation, enabling high-level persistence.

In contrast, LLC genomes displayed overrepresentation of nutrient acquisition systems—notably phosphotransferase systems and ABC transporters—as well as genes involved in motility and chemotaxis, including complete flagellar assembly pathways (Figure 5Q). This signature reflects a competitive, opportunistic strategy oriented toward rapid growth and resource exploitation, yet less compatible with sustained gut colonization.

MLC species exhibited intermediate traits (Figure 5P), with enrichment in sugar metabolism pathways including glycolysis/gluconeogenesis, starch and sucrose metabolism, suggesting metabolic versatility supports moderate-term persistence. These functional distinctions mirror life-history strategies described in classical ecological theory, with convergence across cohorts reinforcing a unifying model: colonization potential is governed by intrinsic ecological adaptation encoded in genome function.

### Intra-species Divergence Drives Clinical Outcomes Across Diseases

Having established that colonization dynamics are governed by ecological attributes, we investigated whether intra-species genetic heterogeneity could explain variable host responses observed following microbiota-based interventions. To this end, we developed a population-scale analytical framework that classifies strains as health-associated or disease-associated based on >98% nucleotide similarity. Strains were categorized as health-associated if they originated from donors or post-FMT responders, and as disease-associated if derived from pre-FMT samples or non-responders.

Applying this approach to our NSCLC cohort, we identified pronounced intra-species divergence in 484 taxa. For instance, *Agathobacter rectalis* (**Figure 6A-C**) and *Bacteroides uniformis* (Supplementary Figure 11A-C) each displayed deep bifurcation into two phylogenetically distinct clades. Clade A strains were significantly enriched in responders, whereas Clade B strains predominated in non-responders (Fisher’s exact test, *p* < 0.001). These associations remained robust after adjusting for relative abundance and inter-individual variation, indicating that strain genotype, rather than species-level abundance, is a key determinant of clinical outcome.

**Figure 6.**
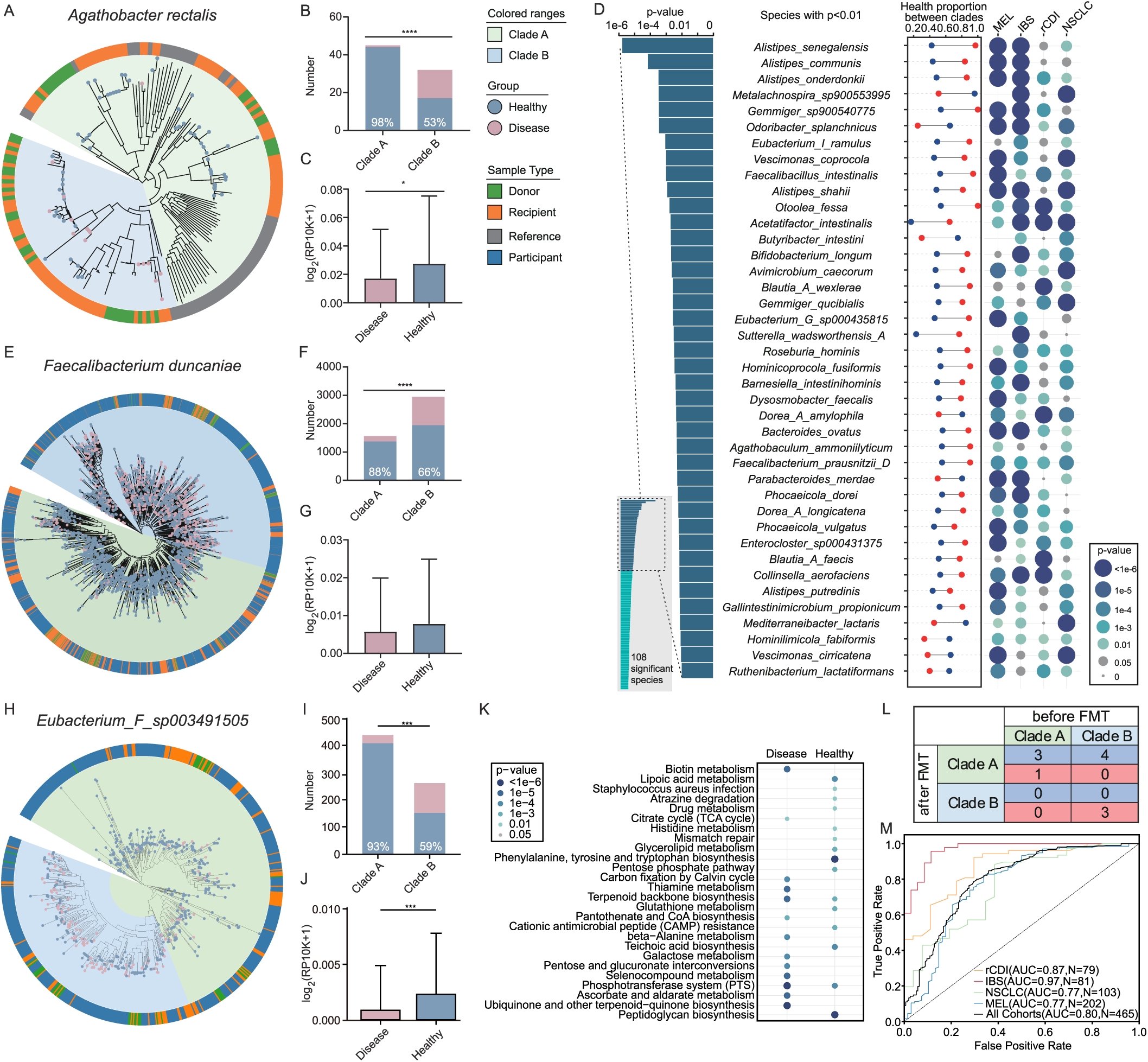
Population-level tracking reveals health- and disease-associated bacterial strain heterogeneity. **A.** Phylogenetic tree of *Agathobacter rectalis* strains from the NSCLC-FMT cohort constructed using ultra-single-copy genes (USCGs). Tree colored by sample type (donor, recipient, reference, participant) with outer ring indicating clade classification (A vs B). **B.** Strain distribution analysis for *A. rectalis* clades showing differential association with health outcomes. Bar plot compares clade proportions between health states of the statistical significance indicated. Numbers indicate proportion of healthy strains. **C.** Relative abundance comparison of *A. rectalis* from healthy and diseased individuals, demonstrating significantly higher abundance in healthy individuals. **D.** Species-level analysis across four FMT cohorts showing subspecies heterogeneity based on p-values for health proportion differences between clades. Bar chart displays p-values (left axis) and dot plot shows health proportions for each clade across cohorts (MEL, IBS, rCDI, NSCLC). Bubble size reflects significance level; gray bubbles indicate non-significant associations (p>0.05). **E.** USCG-based phylogenetic tree of *Faecalibacterium duncaniae* across 8 cohorts with clade designation and sample classification. **F.** Strain distribution of *F. duncaniae* clades across health and disease states, showing preferential association of specific clades with clinical outcomes. **G.** Relative abundance comparison of *F. duncaniae* between health and disease states. **H.** Phylogenetic tree of *Eubacterium F* sp003491505 strains across cohorts with clade classification and sample type annotation. **I.** Strain distribution analysis for *Eubacterium F* sp003491505 clades showing health-disease associations. **J.** Relative abundance analysis of *Eubacterium F* sp003491505 demonstrating differential abundance patterns between healthy and diseased individuals. **K.** KEGG enrichment displaying differentially enriched genes between disease and healthy related clades, with statistical significance thresholds indicated. **L.** Time-series analysis of *A. rectalis* clades in the NSCLC cohort pre- and post-FMT, with SD (blue) and PD (red) outcomes **M.** Receiver operating characteristic (ROC) curve analysis evaluating the discriminatory power of healthy/disease-associated clades in 109 species for health-disease classification across multiple cohorts. For all analyses: SD = successful/healthy outcome; PD = poor/disease outcome. Pre-treatment samples classified as disease; post-treatment responders as healthy; non-responders remain diseased. Statistical significance: *p<0.05, **p<0.01, ***p<0.001.

Consistent patterns of intra-species divergence were observed in the other three FMT cohorts, such as *Bacteroides caccae* in melanoma, *Alistipes onderdonkii* in IBS, and *Parabacteroides distasonis* in rCDI (Supplementary Figure 11D–L). A substantial fraction of divergent species (14%; 66/484) exhibited cross-disease conservation (Supplementary Figure 12). This suggests the existence of universally health-associated strain populations that transcend disease context.

### Universal Convergence of Health-Associated Clades Across Indications

When integrating strain-level data from all four cohorts, we identified 108 species demonstrating statistically significant population segregation, with consistent associations between specific clades and clinical outcomes (Cochran–Mantel–Haenszel test, p < 0.05; Figure 6D). Notably, *Faecalibacterium duncaniae* (Figure 6E–G), *Eubacterium F sp003491505* (Figure 6H–J), *Bifidobacterium adolescentis* (Supplementary Figure 13A–C), *Metalachnospira sp900553995* (Supplementary Figure 13D–F)*, Hominimerdicola aceti* (Supplementary Figure 13D–L) *and Hominimerdicola dorei* (Supplementary Figure 13D–L) exemplified this pattern, each comprising distinct clades with contrasting clinical associations conserved across diseases. These findings underscore the limitations of species-level analysis: in several cases—such as *F. duncaniae, M. sp900553995,* and *H. dorei*—overall abundance did not differ significantly between responders and non-responders, yet strain-resolved analysis revealed clades with opposing effects on treatment response.

Functional profiling elucidated the molecular basis of this divergence. Health-associated strains are generally enriched in biosynthetic and metabolic pathways that support de-novo nutrient synthesis, energy generation, cellular integrity, and detoxification, including phenylalanine/tyrosine/tryptophan biosynthesis, peptidoglycan assembly, and DNA mismatch repair. In contrast, disease-associated strains tend to exhibit functional signatures biased toward stress adaptation, resistance to host defenses, and altered nutrient utilization, exemplified by the phosphotransferase system (PTS) and galactose metabolism pathways (Figrue 6K).

### Strain-Level Trajectories Causally Link Microbial Shifts to Clinical Response

To assess whether strain-level dynamics directly influence therapeutic outcomes, we performed longitudinal tracking of strain populations pre- and post-FMT. In *A. rectalis*, patients who acquired and sustained dominance of Clade A (health-associated) strains post-FMT experienced significantly improved clinical outcomes, while those retaining dominance of Clade B (disease-associated) strains showed poor response (*p* < 0.01) (Figure 6L). Notably, patients who transitioned from Clade B to Clade A post-FMT, indicating successful engraftment of beneficial strains, showed robust and sustained clinical improvement—irrespective of their initial microbial composition. These results argue against a purely subtractive model of pathogen clearance, and instead support a model in which targeted engraftment of functionally beneficial strains drives therapeutic efficacy.

### HSIM Score Predicts Treatment Response and Guides Donor Selection

To operationalize these findings for clinical application, we developed the Healthy Strain in Microbiome (HSIM) score—a metric quantifying the proportion of health-associated strains in each individual’s microbiome. HSIM scores robustly predicted clinical outcomes across all four cohorts, with receiver operating characteristic (ROC) area under the curve (AUC) values of 0.97 for IBS, 0.87 for rCDI, 0.77 for both NSCLC and melanoma, and 0.80 when all diseases were pooled (Figure 6M). These findings demonstrate the cross-cohort predictive utility of strain-level profiling.

Longitudinal analysis revealed that HSIM scores significantly increased following FMT (Supplementary Figure 13M-P), with responders showing larger and more sustained gains than non-responders. Divergence in HSIM trajectories emerged as early as 2–4 weeks post-FMT and remained stable over time. This early and durable separation suggests that HSIM score may serve as a leading indicator of therapeutic response.

### Prioritizing Therapeutic Strains Based on Colonization and Functional Impact

To identify high-priority microbial candidates for therapeutic development, we integrated colonization capacity, ecological persistence, and clinical impact of strain-specific genomic variation. This multi-dimensional screen yielded 39 candidate species characterized by high persistence potential and consistent strain-level associations with outcomes across cohorts, including *Faecalibacterium prausnitzii*, *Faecalibacterium duncaniae* (Table S2).

These taxa represent clinically validated, ecologically stable, and functionally stratified strains that consistently track with favorable clinical trajectories, positioning them as promising leads for next-generation microbiome therapeutics.

## DISCUSSION

This study delivers a fundamental conceptual shift for understanding and predicting the therapeutic efficacy of microbiome interventions by resolving microbial dynamics at the strain level. In the context of advanced PD-L1-negative lung adenocarcinoma (NSCLC)—a notoriously immunotherapy-refractory population—we demonstrate that FMT significantly prolongs progression-free survival when combined with PD-1 blockade. While this finding has direct clinical relevance, its broader significance lies in the universal ecological and genomic principles uncovered through high-resolution metagenomic tracking. Our analysis across over 2,000 samples and multiple disease contexts reveals that the functional identity and ecological fitness of individual microbial strains—not species-level taxonomy—determine therapeutic outcomes. These findings resolve persistent contradictions in the microbiome literature and provide a roadmap for precision microbiota-based therapeutics.

Previous studies have hinted at the importance of strain specificity in shaping host response. Smillie et al.^[13]^ employed whole-genome sequencing and strain-tracking to study rCDI, identifying donor strains that engrafted persistently and correlated with clinical success. However, their analysis was confined to a narrow taxonomic scope, lacked ecological generalization, and was limited to a single disease. Similarly, recent immunotherapy studies^[5, 6]^ identified microbial correlates of response in melanoma cohorts, associating species such as *Akkermansia muciniphila*, *Bifidobacterium longum*, and *Ruminococcus bromii* with positive outcomes. However, these associations remain inconsistent across studies (e.g., *A. muciniphila* was negatively associated with response in colitis and CRC models^[14, 15]^), exposing a persistent reproducibility crisis in microbiome research.

Our findings resolve these contradictions by moving beyond species-level inference. For example, we show that *Faecalibacterium prausnitzii*—often hailed as a “universal health biomarker”—in fact harbors functionally divergent clades, one associated with sustained clinical benefit and the other enriched in non-responders. Species-level analyses, which aggregate these conflicting signals, yield incoherent or null results. Only when analyzed at the strain level do the predictive and mechanistic patterns emerge with clarity and reproducibility.

This principle holds consistently across our multicohort analysis. We applied our ucgMLST framework to four FMT datasets (NSCLC, rCDI, IBS, and melanoma) and four longitudinal cohorts, and observed a highly conserved structure of microbial engraftment and persistence. High-level colonizers—especially from *Bacteroidaceae* and *Rikenellaceae*—exhibited high FMT rates and ecological resilience, regardless of disease context or donor-recipient pairings. Conversely, low-level colonizers from *Oscillospiraceae* and *Ruminococcaceae* rarely established durable niches. These findings point to a unifying ecological architecture underlying FMT outcomes—one shaped not by host condition or donor-recipient identity, but by genome-encoded life history strategies of the microbes themselves.

Critically, we show that these ecological traits correlate directly with clinical outcomes. In NSCLC patients, responders consistently exhibited higher rates of successful strain engraftment, defined by sustained presence of donor strains with low pre-treatment similarity and high ecological fitness. Responders showed not only elevated proportions of high-level colonizers but also significantly lower similarity to their pre-FMT microbiota, indicating true ecological replacement rather than transient enrichment. Moreover, we observed a bimodal distribution in strain sharing between donors and recipients, allowing us to identify a discrete engraftment threshold (12.5%) that distinguished successful from unsuccessful colonization events. This distribution was predictive of treatment response, suggesting that FMT efficacy may not be stochastic but rather governed by deterministic colonization barriers that can be measured and potentially manipulated. This echoes results from Chen-Liaw et al.^[26]^, who recently showed that strain richness and compatibility, rather than mere abundance or alpha diversity, predict engraftment and host benefit.

These results affirm and extend observations from previous studies^[24, 30]^, which emphasized host-microbiome compatibility but lacked a framework for cross-cohort generalizability. By classifying strains into health- or disease-associated populations based on clade-level phylogenetic structure, we show that these strain-clinical associations are conserved and reproducible across diverse host states, geographies, and interventions. To translate these findings into a clinically actionable tool, we developed the HSIM score, which quantifies the proportion of health-associated strains within an individual’s gut microbiome. HSIM scores robustly predicted clinical outcomes, with area under the ROC curve values of 0.97 in IBS, 0.87 in rCDI, and 0.77 in both NSCLC and melanoma. Longitudinal analysis revealed that HSIM scores diverged between responders and non-responders as early as two weeks post-FMT and remained stable over time, suggesting that this metric could serve as a leading indicator of therapeutic response. Furthermore, recipients who received FMT from donors with high HSIM scores exhibited significantly greater increases in post-FMT HSIM values and higher probability of clinical benefit, underscoring the utility of HSIM for donor selection and personalized treatment stratification.

Importantly, our findings provide a mechanistic explanation for these predictive associations. HLCs consistently encoded genomic features linked to mucosal colonization, immune evasion, and anaerobic energy metabolism, including genes involved in amino sugar utilization, extracellular polysaccharide biosynthesis, and phosphotransferase systems. These traits likely confer selective advantages in the inflamed, nutrient-limited gut environments characteristic of many disease states, and may also promote regulatory immune interactions with the host. In contrast, LLCs were enriched for flagellar biosynthesis, chemotaxis, and mobile genetic elements—features commonly associated with ecological opportunism, instability, and inflammation. These results support a model in which host-compatible, K-selected strains engraft durably and mediate therapeutic benefit, while r-selected strains drive instability or elicit deleterious immune responses. This evolutionary lens offers a coherent framework for understanding how microbial ecology and function converge to shape host outcomes.

While the conceptual and analytic advances presented here resolve key contradictions in the field, they also open several new avenues of investigation. A key limitation of our study is the modest size of the NSCLC cohort and the absence of a randomized control arm. To address this, we benchmarked clinical outcomes against rigorously matched real-world controls obtained from a city-wide digital registry aggregating longitudinal electronic health records across multiple hospitals. These data were automatically extracted, fully de-identified, and filtered to ensure treatment consistency, minimizing the risk of selection or reporting bias. Nonetheless, real-world data cannot fully substitute for the internal validity provided by randomization and blinding. Future trials should incorporate prospective randomized arms and stratified donor selection based on HSIM or other strain-resolved metrics to validate these associations with greater statistical power and clinical generalizability.

Additionally, while our strain tracking framework captures a wide range of genomic variation, extremely low-abundance or recombinant strains may evade detection, particularly in cases of horizontal gene transfer or genomic rearrangement. Finally, although our data support strong associations between strain engraftment and clinical benefit, definitive causal links require mechanistic validation. Gnotobiotic models colonized with health- and disease-associated clades from our priority species—such as *Eubacterium* F sp003491505—could clarify functional roles and define host pathways targeted by microbial metabolites or surface structures.

From a translational perspective, this study sets the stage for a new generation of microbiome therapeutics. By prioritizing strains that are not only ecologically robust but also functionally aligned with clinical benefit, we move beyond artisanal donor selection and toward rational design of defined microbial consortia. The 39 candidate species identified here represent lead taxa for development as live biotherapeutic products (LBPs), each validated across multiple cohorts and carrying conserved functional signatures of benefit. Moving forward, these strains can be isolated, cultivated under GMP conditions, and assembled into synthetic communities optimized for colonization, stability, and immunomodulatory potential. Importantly, our findings suggest that such consortia need not recapitulate full microbial diversity; rather, therapeutic efficacy may depend on the engraftment of a few well-characterized, high-fitness strains with known host compatibility.

In conclusion, this work redefines the foundation of microbiome therapeutics by demonstrating that strain-level genetic identity and ecological adaptation are the primary determinants of FMT efficacy. By integrating multi-cohort strain tracking, functional genomics, and longitudinal clinical data, we provide a unifying framework that resolves previous contradictions, identifies conserved predictors of response, and enables precision engineering of microbial interventions. As the field moves toward clinical deployment of microbiota-based therapies, the principles and tools described here will be essential for ensuring reproducibility, scalability, and therapeutic success.

## METHODS

### Study design and cohort enrollment

We analyzed four independent FMT cohorts representing diverse disease contexts:

1. **Lung adenocarcinoma cohort:** Patients with advanced lung adenocarcinoma (stage IIIB/IIIC/IV) and negative or low (<1%) PD-L1 expression were enrolled between December 2021 and June 2023 (Clinical Trial Registration: ChiCTR2300076829). Patients received standard first-line therapy (platinum-based chemotherapy plus pembrolizumab) combined with oral FMT capsules from carefully screened healthy donors.
2. **Irritable bowel syndrome cohort:** Patients with moderate-to-severe IBS (n=28) received FMT via colonoscopy from healthy donors, with longitudinal sampling over 24 weeks.
3. **Recurrent C. difficile infection cohort:** Patients with recurrent CDI (n=19) received FMT via colonoscopy, with follow-up samples collected at 2, 8, and 24 weeks post-treatment.
4. **Melanoma cohort:** Patients with advanced melanoma (n=15) receiving anti-PD-1 therapy were administered FMT capsules from donors selected based on previous response to immunotherapy.

All studies were approved by the respective Institutional Review Boards, and all participants provided written informed consent.

### Donor screening and FMT preparation

Across all cohorts, healthy donors underwent rigorous screening according to FDA guidelines for FMT, including testing for bloodborne pathogens, enteric pathogens, multi-drug resistant organisms, and parasites. Donors maintained controlled diets during the collection period, avoiding potential immunomodulatory foods.

Fecal material was processed under anaerobic conditions within 6 hours of collection. Briefly, stool was homogenized in sterile saline containing 12.5% glycerol, filtered through a 330 μm filter, and concentrated by centrifugation. The resulting material was administered via the protocol-specific route (oral capsules, colonoscopy, or duodenal infusion).

### Metagenomic sequencing and quality control

DNA extraction from fecal samples was performed using the HiPure Bacterial DNA kit (Magen, D3146) according to the manufacturer’s protocol. Library preparation used the VAHTS Universal DNA Library Prep kit (Vazyme, ND607) with 300 bp inserts. Quality control included Qubit quantification and Agilent 2100 fragment analysis.

Sequencing was performed on an Illumina NovaSeq 6000 platform using the NovaSeq 6000 S4 Reagent Kit v1.5, generating 150 bp paired-end reads with an average depth of 6 Gb per sample. Raw reads underwent quality control using EToKi (v1.3) to remove low-quality sequences (Q<20) and adapter contamination. Host-derived reads were removed using BBduk against the human reference genome (GCF_000001405.40).

### Real-world data extraction

This study was designed as a single-arm trial employing real-world evidence (RWE) as an external control. Because the inclusion and exclusion criteria were applied to registry-derived cases from the same calendar period, the external comparator qualifies as a concurrent external control. Environmental comparability was ensured by aligning the historical context, diagnostic modalities, disease taxonomy, and treatment technologies between the experimental and control cohorts. To address therapeutic comparability, control-patient selection incorporated rigorous adjustment for baseline characteristics, intervention details, and potential confounders, as detailed below.

Patient selection was performed in four steps. First, the entire institutional repository—including outpatient encounters, admission notes, progress records, discharge summaries, and front-page medical charts—was screened using ICD-10 code C34 to establish a master roster of lung-cancer patients. Second, this roster was queried with the keywords “non-small-cell adenocarcinoma,” “advanced/metastatic,” and “PD-1 monoclonal antibody/immunotherapy,” yielding 1,479 candidate cases. Third, patients were excluded in a stepwise manner if they had TNM stage I–II disease, PD-L1 TPS ≥1% or unknown PD-L1 status, incomplete combination therapy, regimens lacking platinum-based chemotherapy, concomitant targeted agents, or complex comorbidities such as synchronous malignancies; records with logically inconsistent survival data (OS < PFS) or missing key variables were also removed. Finally, the date of progression was verified against the most recent medical report, and the date of death was cross-validated with official death certificates, resulting in 23 patients eligible for the final analysis.

### Identification of the ultra-conserved single-copy genes

To identify a set of orthologous genes universally present across the tree of life, including bacteria, we first exacted all 573 orthologous genes that have previously been identified by BUSCO ^2^ for evaluation of the completeness of assembled genomes. We did not use their genes directly, because (1) BUSCO established a set of core genes for each taxonomic group, which was not available for metagenomic reads, and (2) they evaluated the presence of core genes based on only 364-2783 genomes from each kingdom, which represent only a small fraction of existing genetic diversity.

We screened the orthologous genes independently using a curated database derived from GenBank. To this end, a total of 556,157 genomes were downloaded from genbank (as of June 2024), consisting of 332,209 from bacteria, 184,818 from virus, 20,155 from eukaryote and 18,975 from archaea, respectively. We estimated the pair-wise genetic distances of the genomes using BinDash, and subdivided them into 116,687 single-linkage clusters of <0.02 genetic distance. One representative sequence was selected for each cluster and used to determine the presences of the 573 orthologous genes searching the HMM profile on all potential coding frames in the sequences. Only genes that were present in single copies in ≥ 70% of the genomes from at least one of the bacteria, archaea, fungi, protozoa, and invertebrates are kept, resulting in a total of 480 genes (https://github.com/Zhou-lab-SUDA/StrTrak/tree/main/uscg_profile), which were designated as ultra-conserved single-copy genes (USCGs).

### Development and implementation of ucgMLST

Using the 480 USCGs, the ucgMLST pipeline consists of two modules: the geneProfile module, which reconstruct metagenome-assembled core genotypes (MACGs) from metagenomic data via read mapping, with optimized parameters (minimum coverage 1×, sequence identity threshold 0.001) for low-abundance strain detection, and the genePhylo module, which generates maximum likelihood phylogenetic trees for evolutionary analysis using IQTree v1.6.12 with automatic model selection.

To validate ucgMLST, we created simulated metagenomic datasets with known strain compositions at various sequencing depths (0.1-10×). Performance was compared against StrainPhlAn and MIDAS using sensitivity, specificity, and accuracy metrics. Real-world validation used public datasets from well-characterized transmission events.

### Performance Benchmarking for ucgMLST

A benchmark dataset of 1 million metagenomic reads was simulated from 29 *Streptococcus* genomes to test species discrimination capabilities of metagenomic pipelines. ucgMLST accurately identified all 29 species with no false positives, outperforming Kraken2 and MetaPhlAn4 in terms of both species identification accuracy and correlation with simulated relative abundances (Figure 3D, Table S3). We spotted a substantially better correlation between the relative abundances of the *Streptococcus species* in the simulation and the predictions by ucgMLST (R^2^=0.98), compared to only 0.83 and 0.90 for predictions from Kraken2 and MetaPhlAn4, respectively (supplementary Figure 7A-C). We also observed high correlations between the pairwise genetic distances calculated based on the USCGs and the core genomes, with a R^2^ of 0.95 (Figure 3E), suggesting sufficient resolution for strain tracing. Using real metagenomic data in our NSCLC cohort, ucgMLST managed to reconstruct strains of low quality with ∼ 0.02X coverage, or high-quality strains with ∼1X coverage, resulting in more strains than StrainPhlAn in all species (**Figure 3F-G**).

We compared the utility of USCGs with whole genomes for *Staphylococcus aureus* infection outbreak identification and found that USCGs can accurately reconstruct the same phylogenetic clusters as whole genomes (supplementary Figure 8D-G). Clinical validation confirmed its utility again: ucgMLST reliably tracked probiotic strain engraftment across multiple time points in longitudinal samples (Figure 3H), distinguishing individual-specific colonization events.

### FMT rate and persistence rate calculations

For each species, FMT rate was calculated as the proportion of donor strains that successfully established in recipients following FMT. A strain was considered successfully transferred if the recipient strain after FMT clustered with the donor strain in the phylogenetic tree with a branch length distance below a defined threshold (0.001 substitutions per site).

Persistence rate was calculated as the proportion of time points at which a strain was detected in longitudinal samples from the same individual. Species were classified as high-level colonizers (LLC, persistence rate ≥66%), mid-level colonizers (MLC, 34-66%), or low-level colonizers species (LLC, ≤34%).

## Statistical analysis

Alpha diversity indices (Shannon, Simpson) were calculated using the Vegan package (v2.6-2) in R (v4.2.1). Beta diversity used Bray-Curtis dissimilarity with significance assessed by PERMANOVA. Differential abundance analysis used LEfSe (v1.1.2) with LDA score ≥3 and p<0.05 as thresholds.

For survival analysis, Kaplan-Meier curves were compared using the log-rank test, with hazard ratios calculated using Cox proportional hazards regression. Time-series analysis of strain dynamics used linear mixed-effects models to account for repeated measures, with p-values adjusted for multiple testing using the Benjamini-Hochberg method.

Strain-specific distribution analysis used pairwise genetic distances between strains, with Fisher’s exact test to identify significant associations with clinical states. All statistical analyses were performed in R (v4.2.1), with p<0.05 considered statistically significant unless otherwise specified.

### Functional genomic analysis

Reads were quality-trimmed (EToKi prepare), assembled (SPAdes v3.13 via EToKi assemble), and annotated (PROKKA v1.14.6, eggnog-mapper v2). Multiple sequence alignments were generated (EToKi align), and maximum likelihood phylogenetic trees were constructed (IQTree v1.6.12 via EToKi phylo). Functional pathway enrichment analysis used clusterProfiler in R with GO and KEGG annotations.

### Data and code availability

Raw sequencing data will be deposited in the Genome Sequence Archive (GSA) under BioProject accession [PRJCA044560]. The ucgMLST code, reference databases, and analysis scripts will be available on GitHub [https://github.com/Zhou-lab-SUDA/ucgMLST]. Additional data supporting the findings of this study are available from the corresponding author upon reasonable request.

## ACKNOWLEDGMENTS

The project was primarily supported by the National Natural Science Foundation of China (32170003, 32370099), the Provincial-level Talent Program for National Center of Technology Innovation for Biopharmaceuticals (NCTIB2024JS0101), the Natural Science Foundation of Jiangsu Province (BK20243008), the Suzhou Top-Notch Talent Groups (ZXD2022003). We thank Ms. Yusu Xia for her assistance with figure and graphic preparation.

---

**Supplementary Figure 1.**
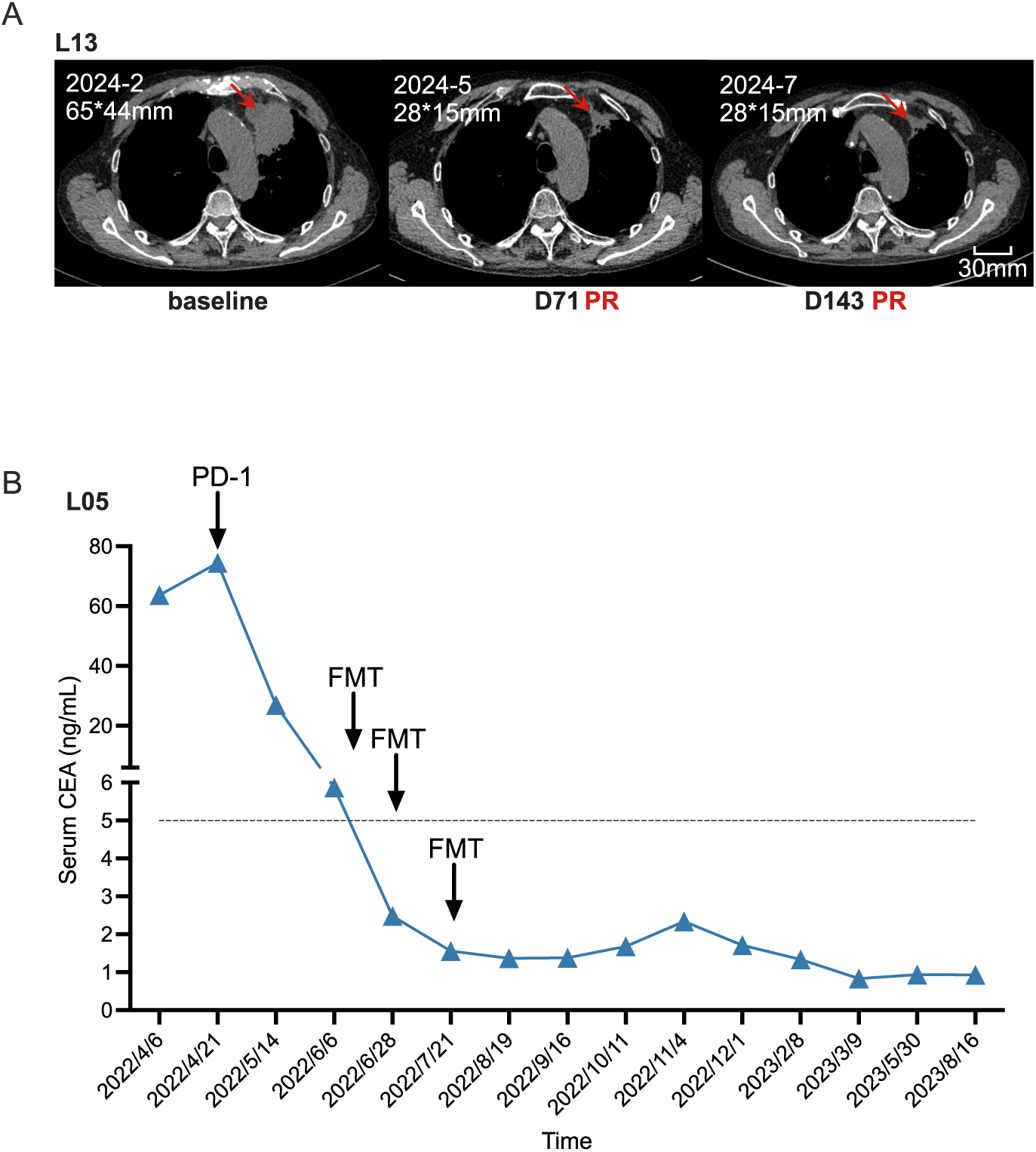
Response to combination therapy in lung cancer. **A.** Contrast-enhanced thoracic CT images of a lung cancer patient (L13) before (baseline) and after treatment (2 months and 5 months after therapy). Red arrows indicate the target lesion. **B.** Concentration of serum CEA of a patient (L05) is dramatically decreased along with the combination treatment.

**Supplementary Figure 2.**
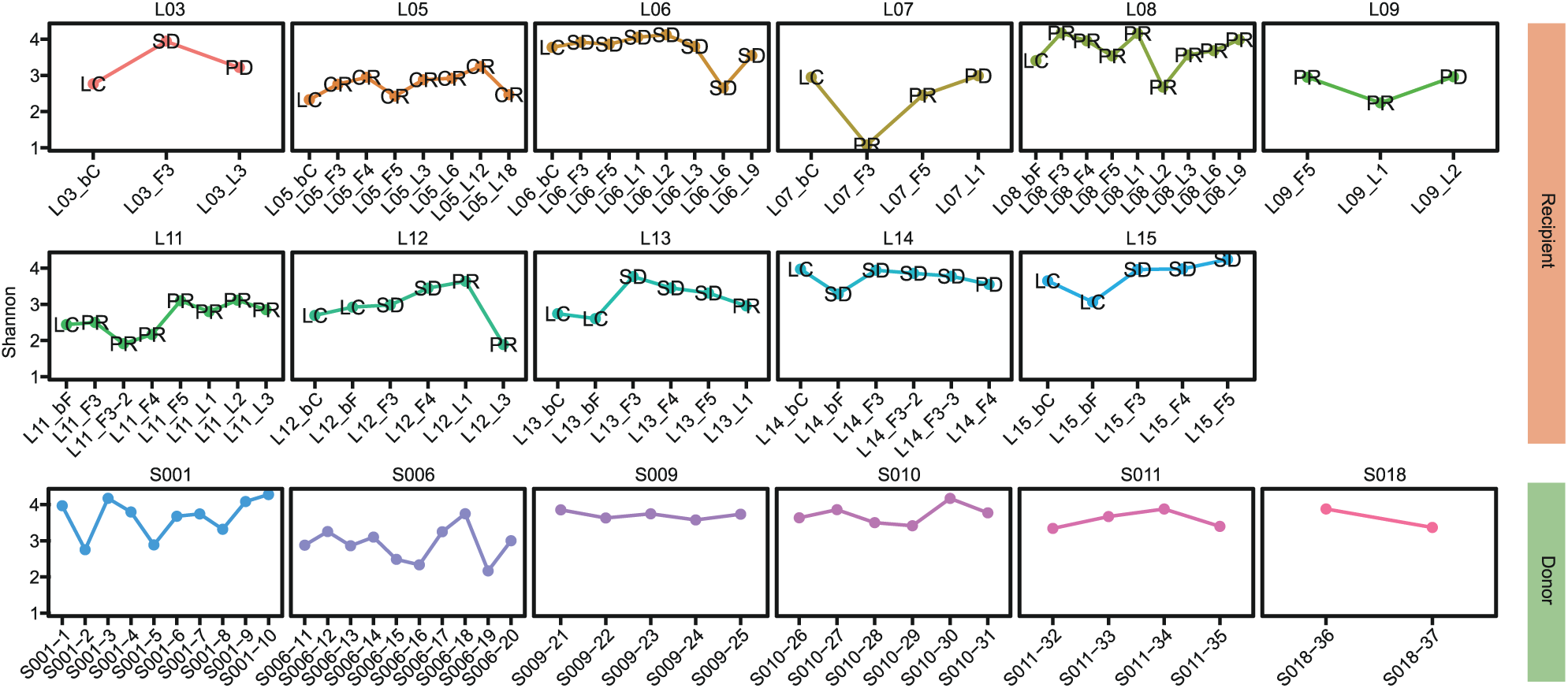
α-Diversity in the NSCLC-FMT Cohort. Colors distinguish donors (green) and recipients (orange). Recipient samples are annotated with efficacy evaluations: NSCLC (pre-FMT), SD (stable disease), PR/CR (partial/complete response), PD (disease progression). The x-axis arranges samples chronologically.

**Supplementary Figure 3.**
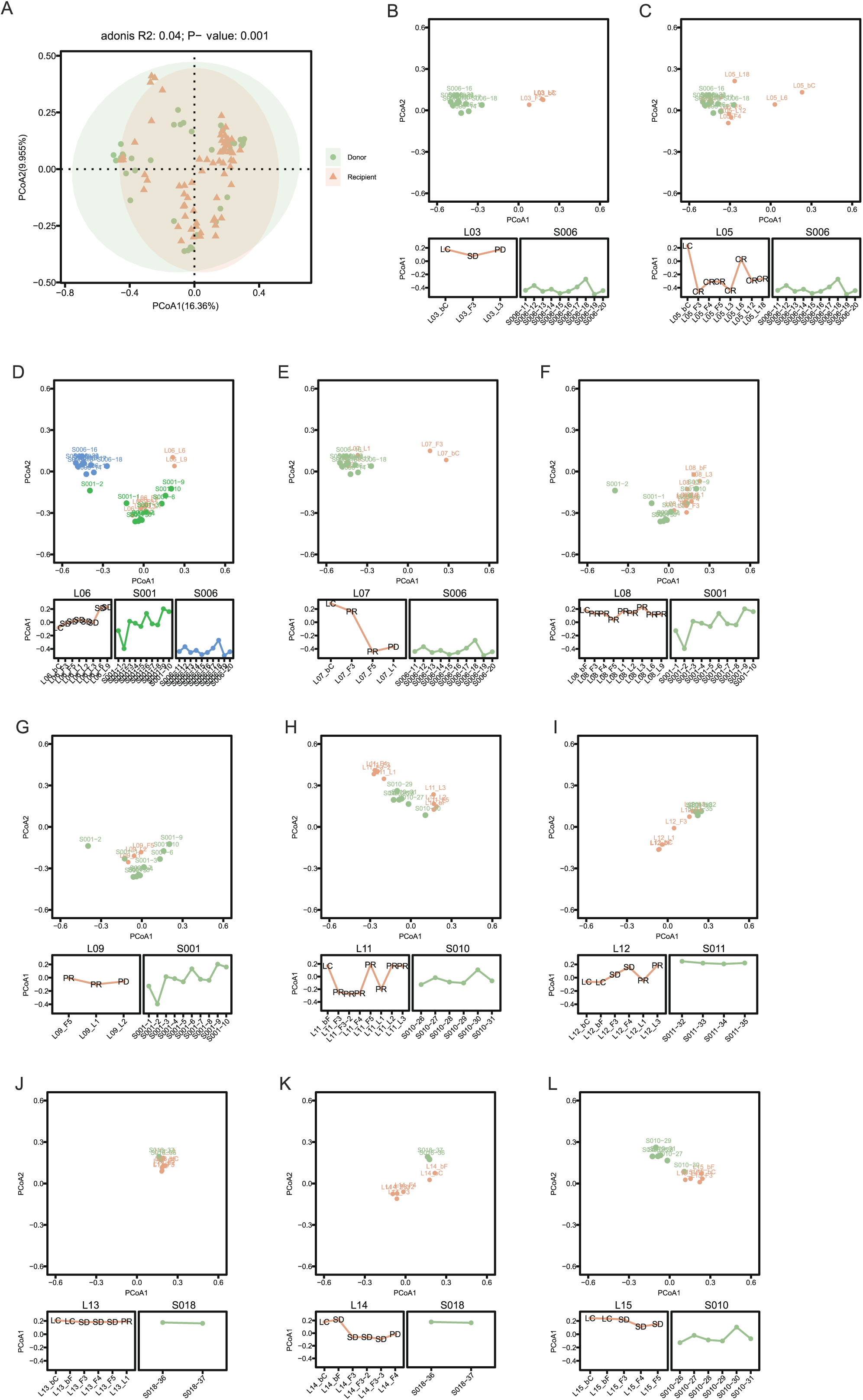
β-Diversity in the NSCLC-FMT Cohort. **A.** PCoA showing significant β-diversity differences between donors and recipients (including all pre- and post-FMT samples; adonis R²=0.04, p=0.001). **B–L.** Dynamic β-diversity changes pre- and post-FMT for each recipient. Some recipients (C, E, H, I) show convergence toward donor β-diversity post-FMT, while others (B, F, G) show no significant change.

**Supplementary Figure 4.**
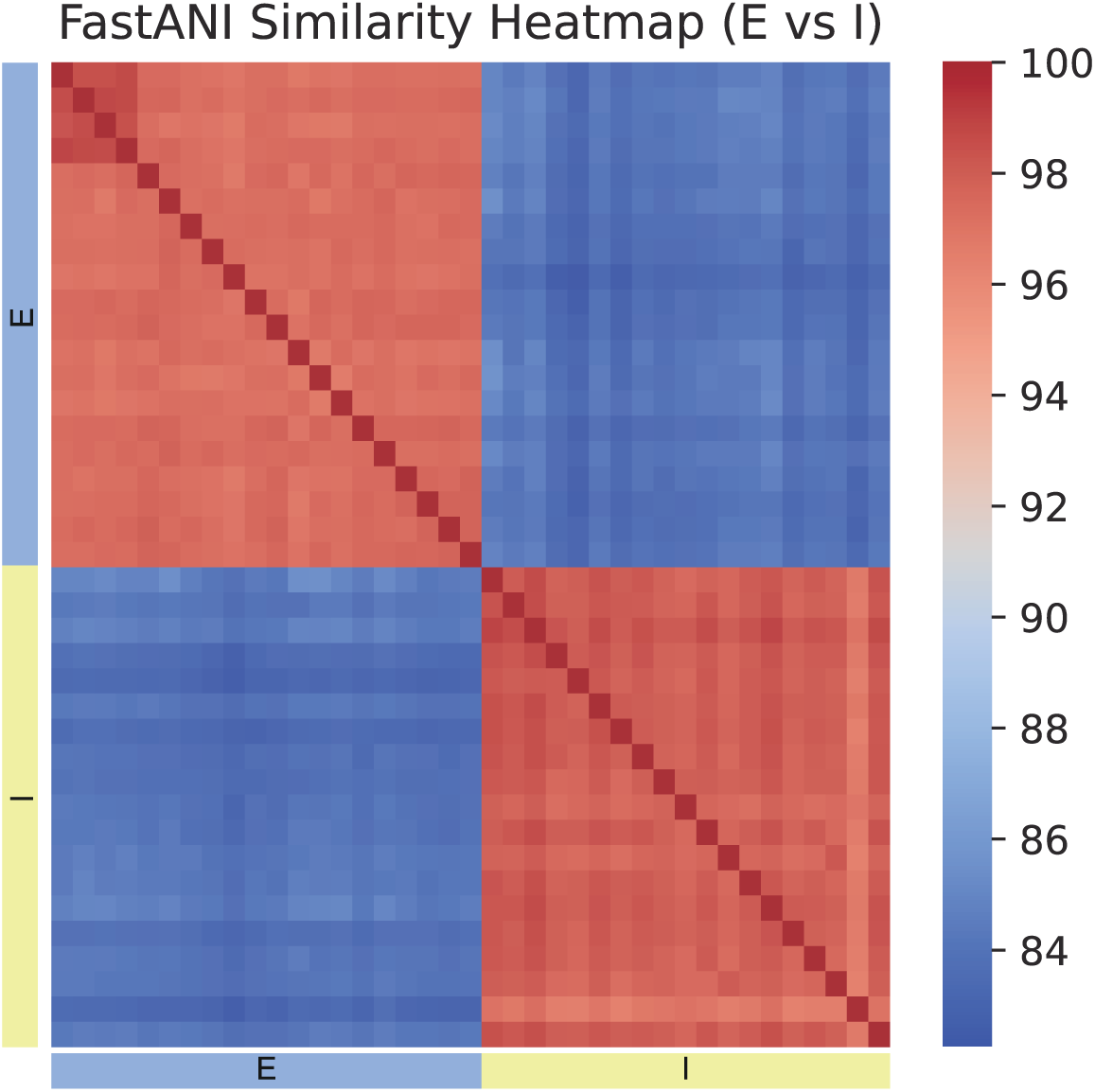
Genetic Similarity Between *F. prausnitzii-E* and *F. prausnitzii-I*

**Supplementary Figure 5.**
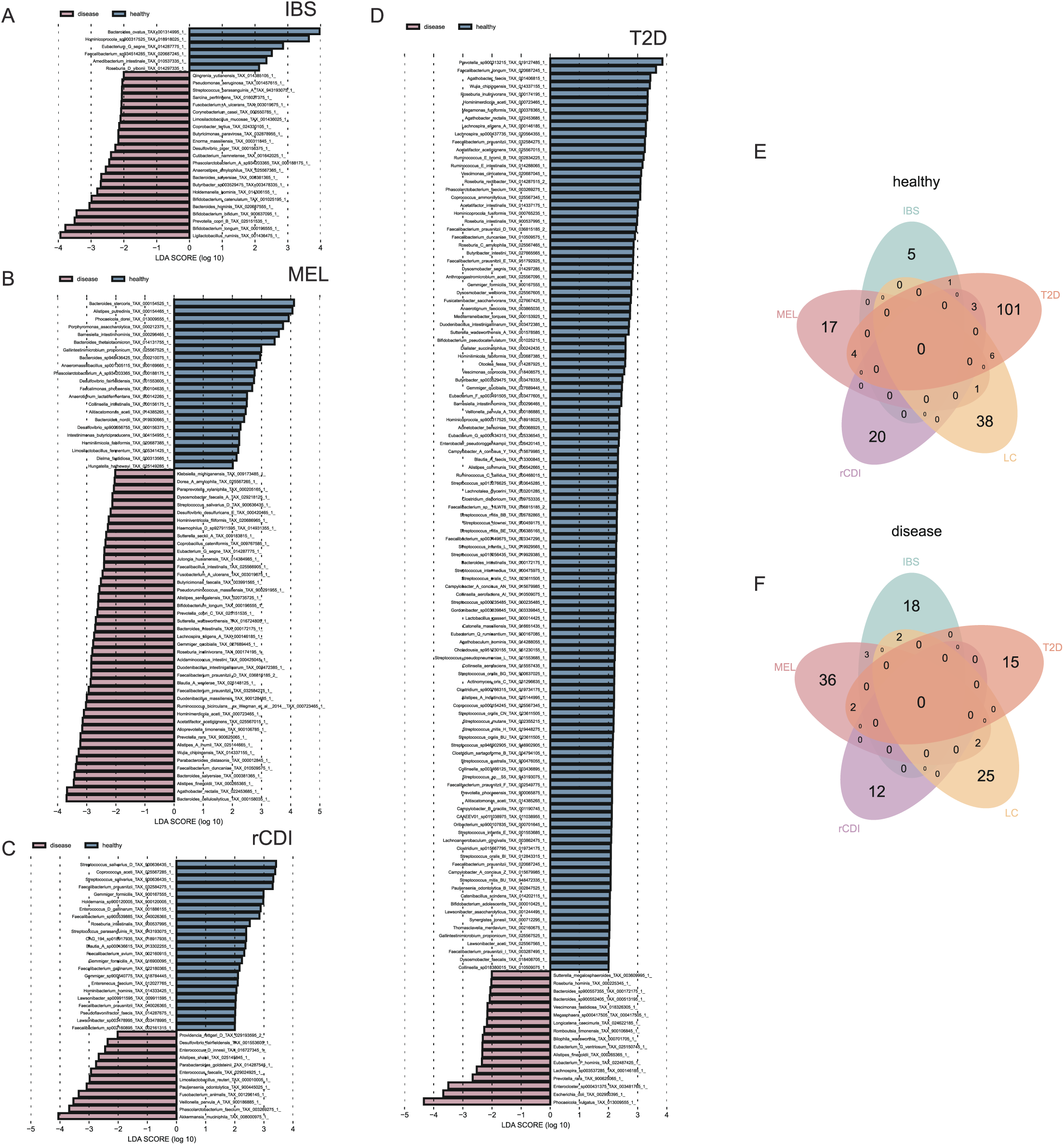
Microbial Enrichment Across Cohorts. **A-D.** LEfSe analysis of microbial enrichment in health versus disease states in the IBS cohort (A), MEL cohor t(B), rCDI cohort (C), T2D cohort (D). **E-F.** Venn diagram of health-associated (E) or disease-associated (F) microbial intersections across cohorts.

**Supplementary Figure 6.**
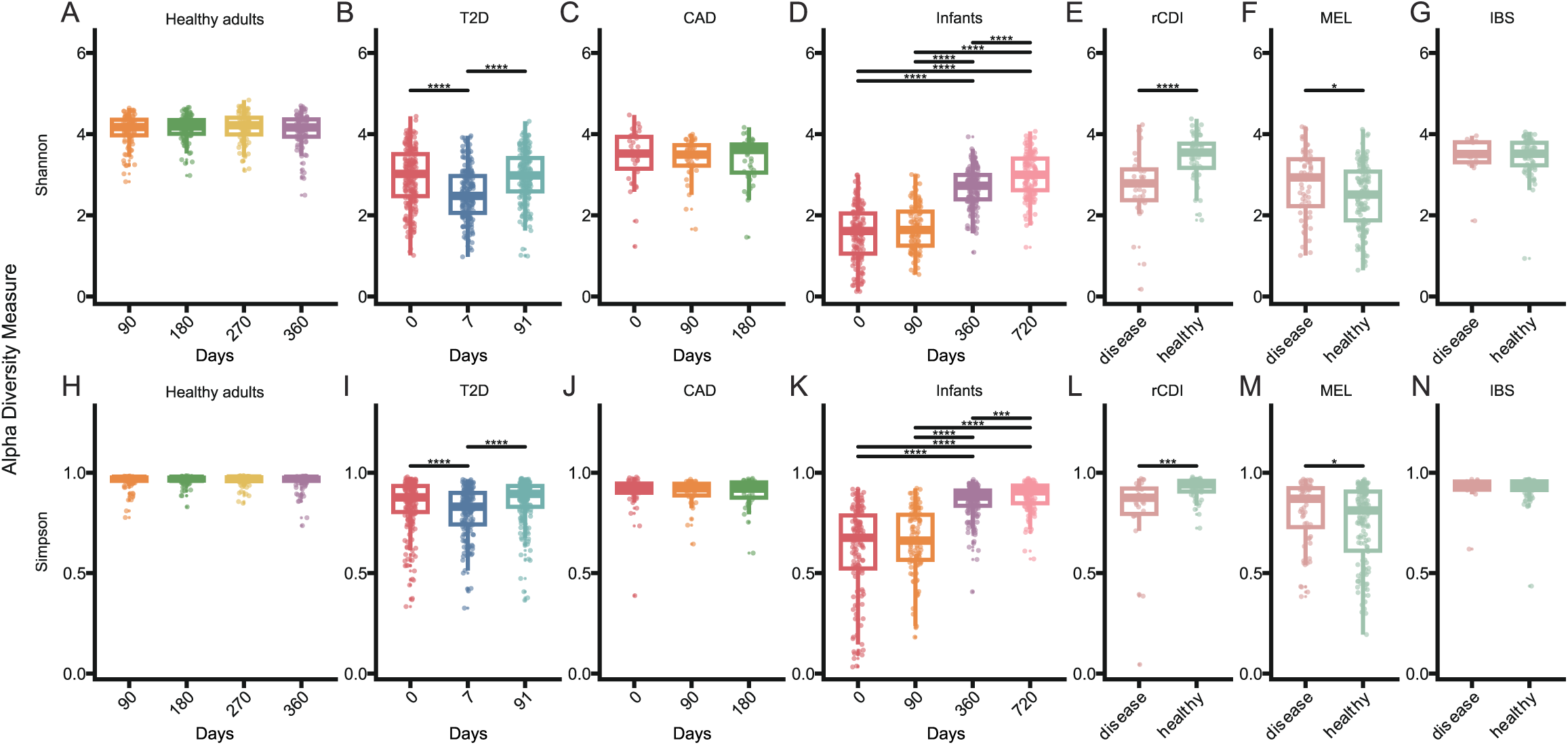
α-Diversity in Public Cohorts

**Supplementary Figure 7.**
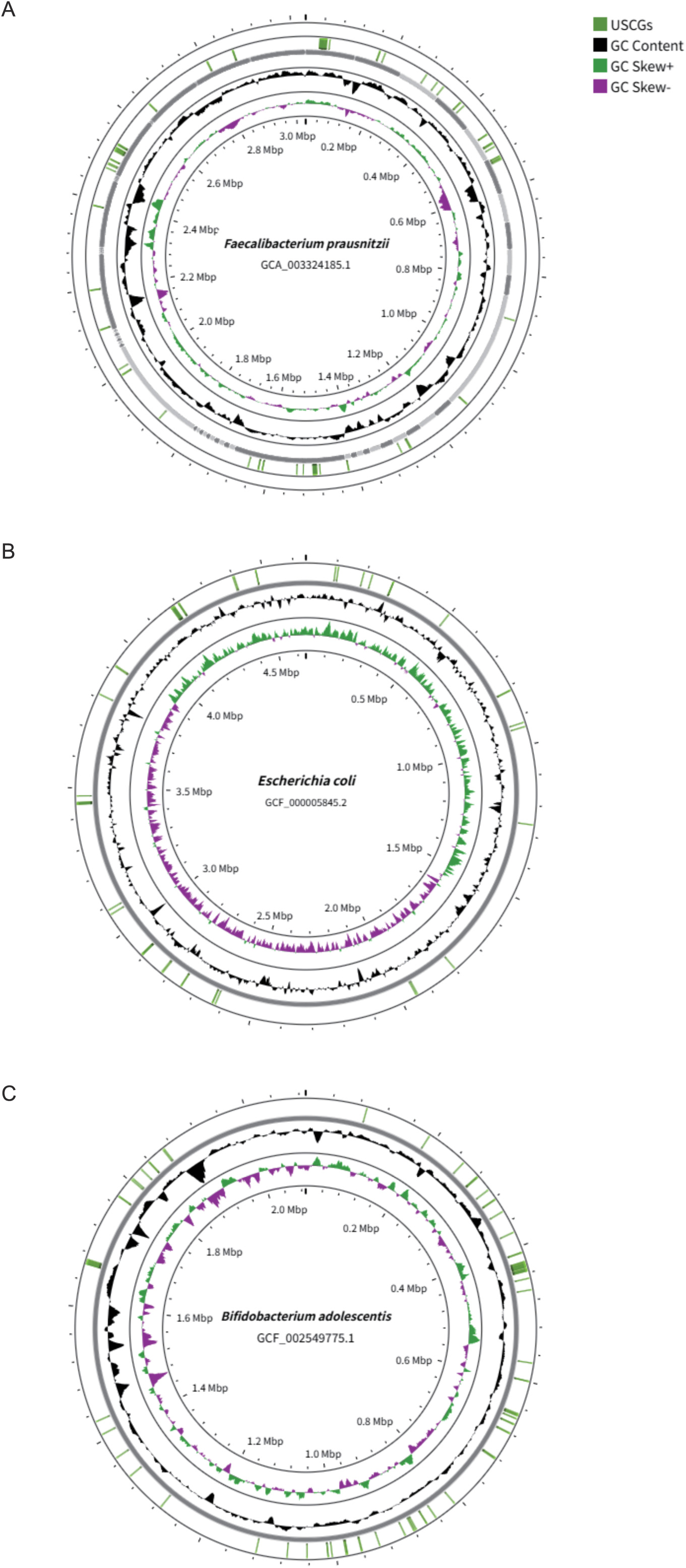
Distribution of USCGs on the genome of Faecalibacterium prausnitzii, Escherichia coli, Bifidobacterium adolescentis.

**Supplementary Figure 8.**
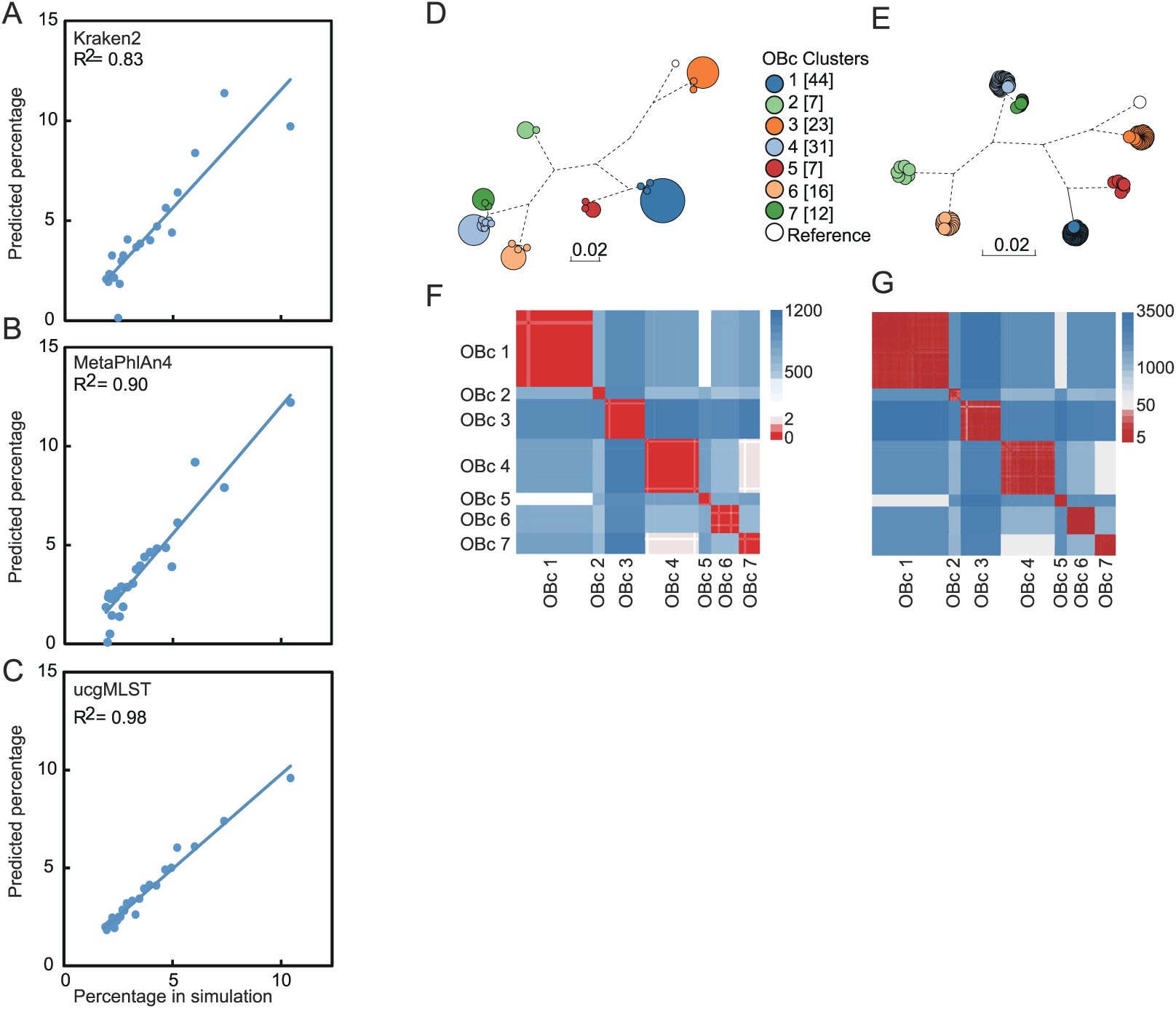
Performance Benchmarking of ucgMLST.

**Supplementary Figure 9.**
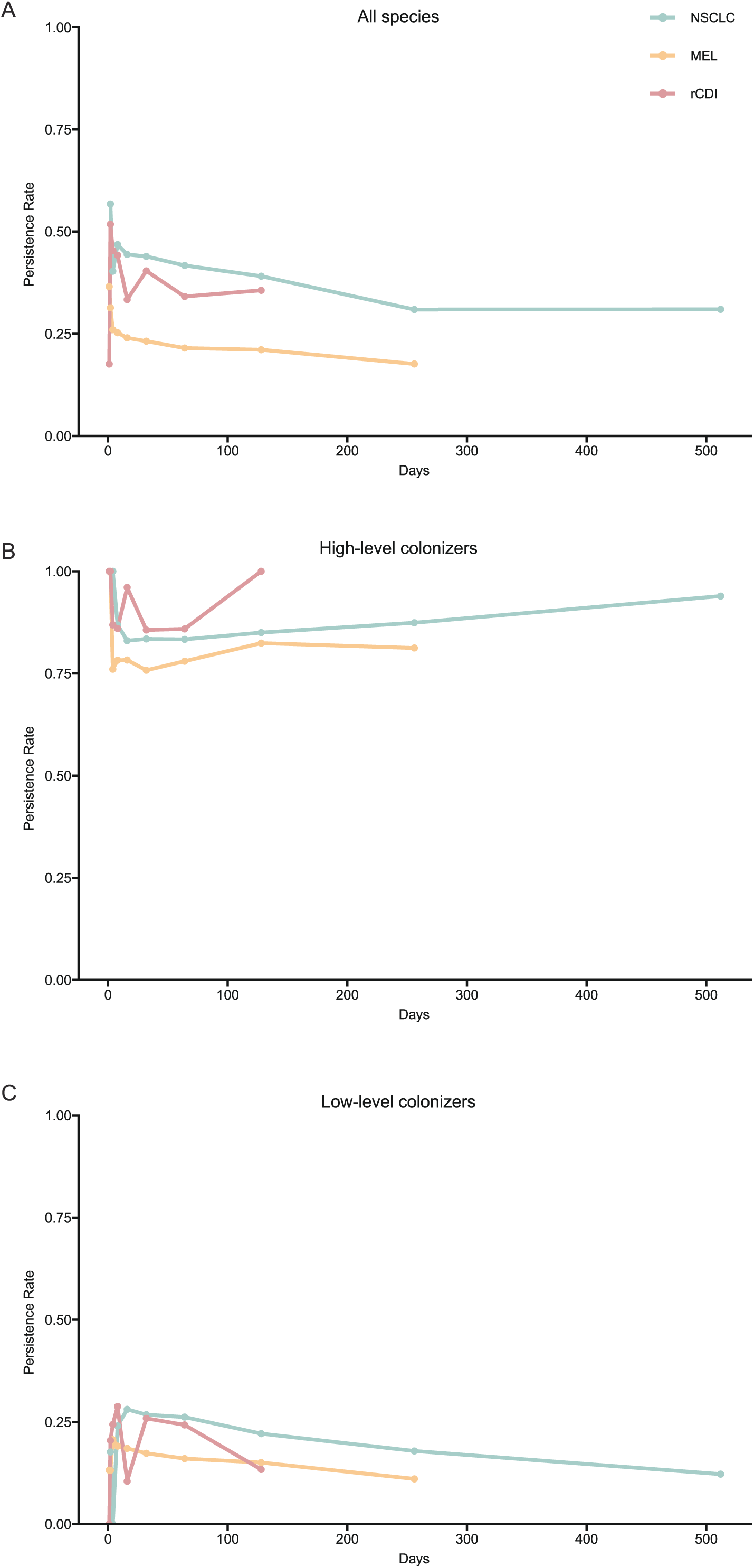
The persistence rate of species changes over time.

**Supplementary Figure 10.**
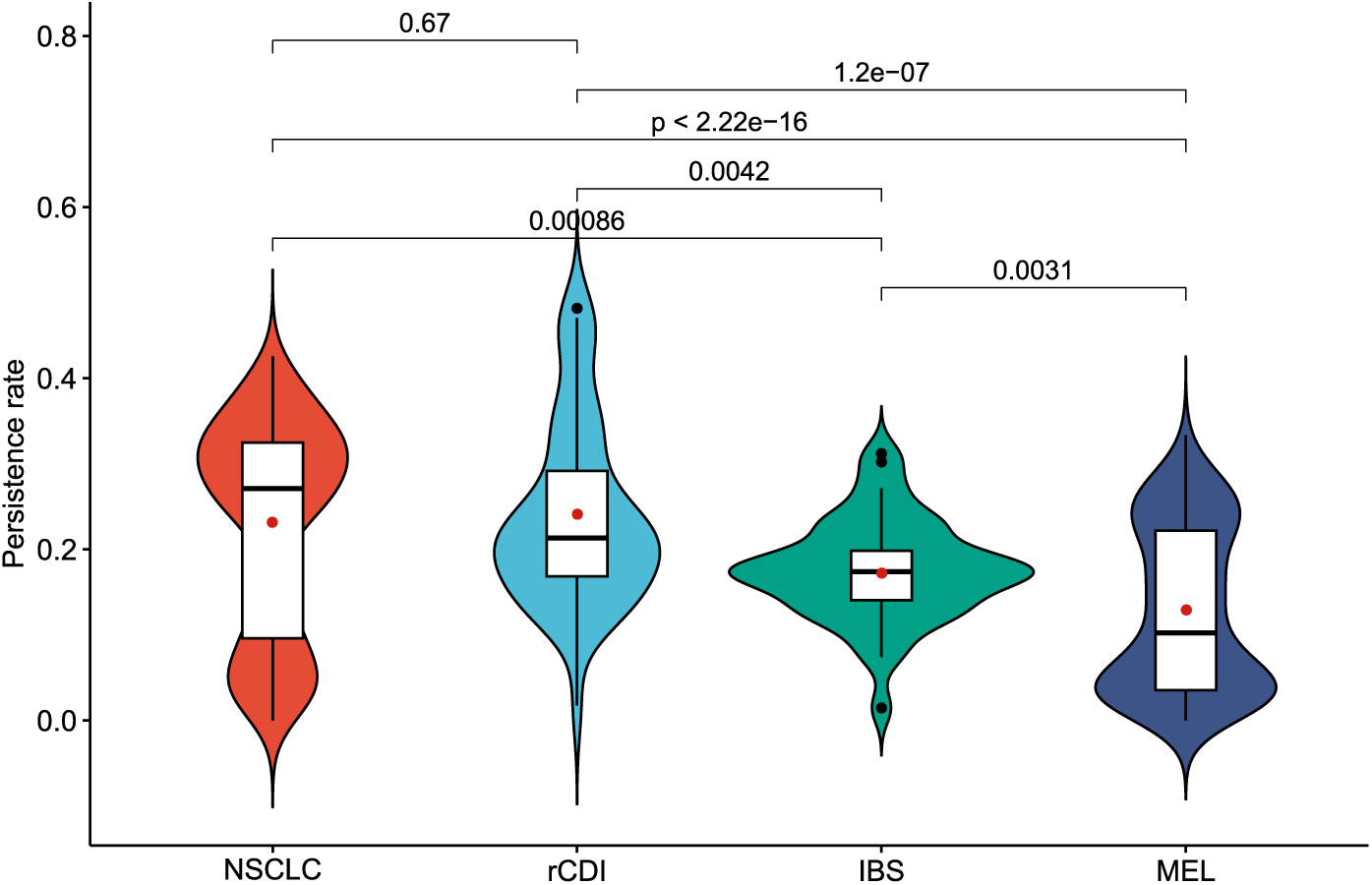
Comparation of average persistence rate of cohorts.

**Supplementary Figure 11.**
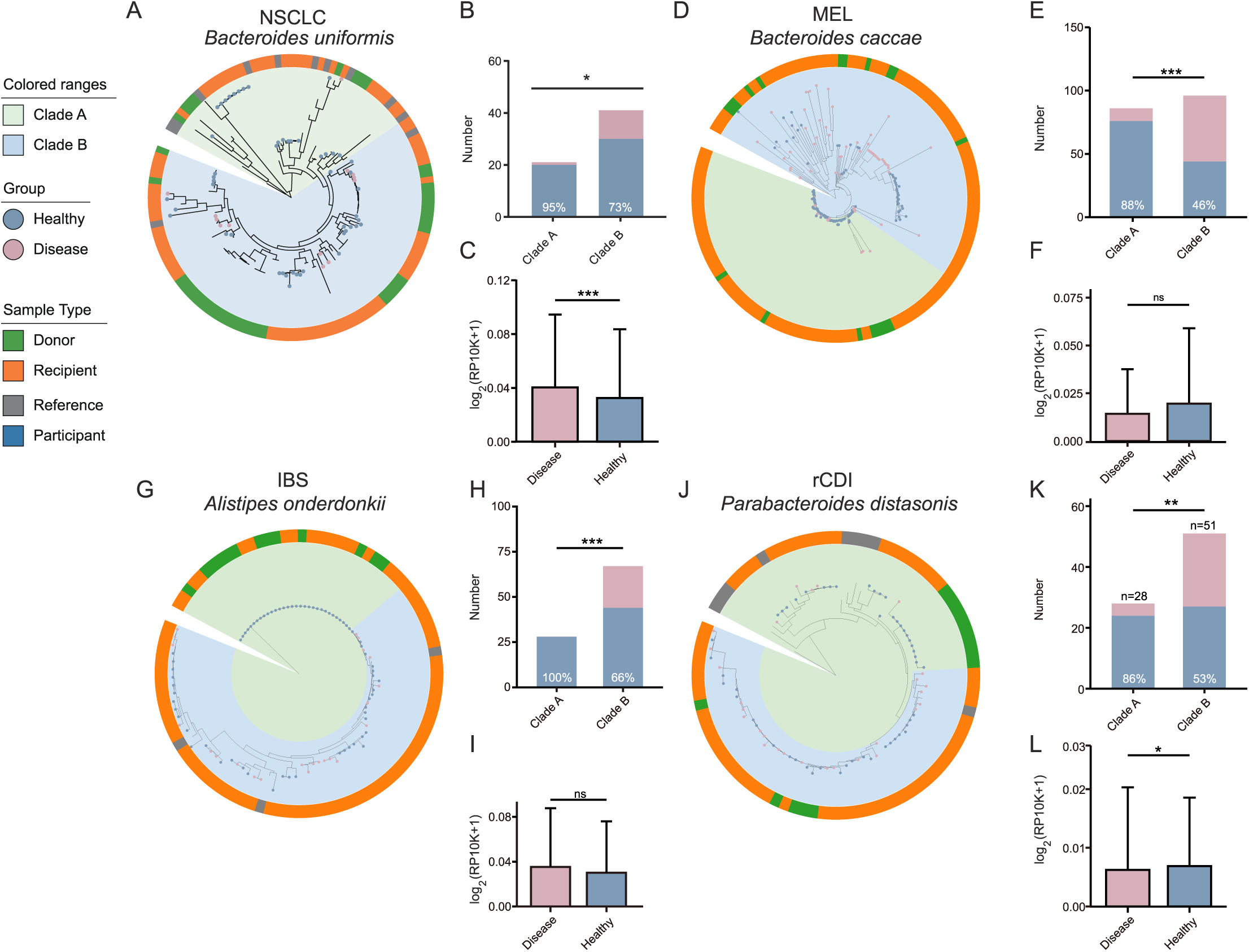
Phylogenetic Analyses of species from single corhort. **A–L.** USCG-based phylogenetic analyses of (A-C) *Bacteroides uniformis* in the NSCLC cohort, (D-F) *Bacteroides caccae* in the MEL cohort, (G-I) *Alistipes onderdonkii* in the IBS cohort and (J-L) *Parabacteroides distasonis* in the rCDI cohort.

**Supplementary Figure 12.**
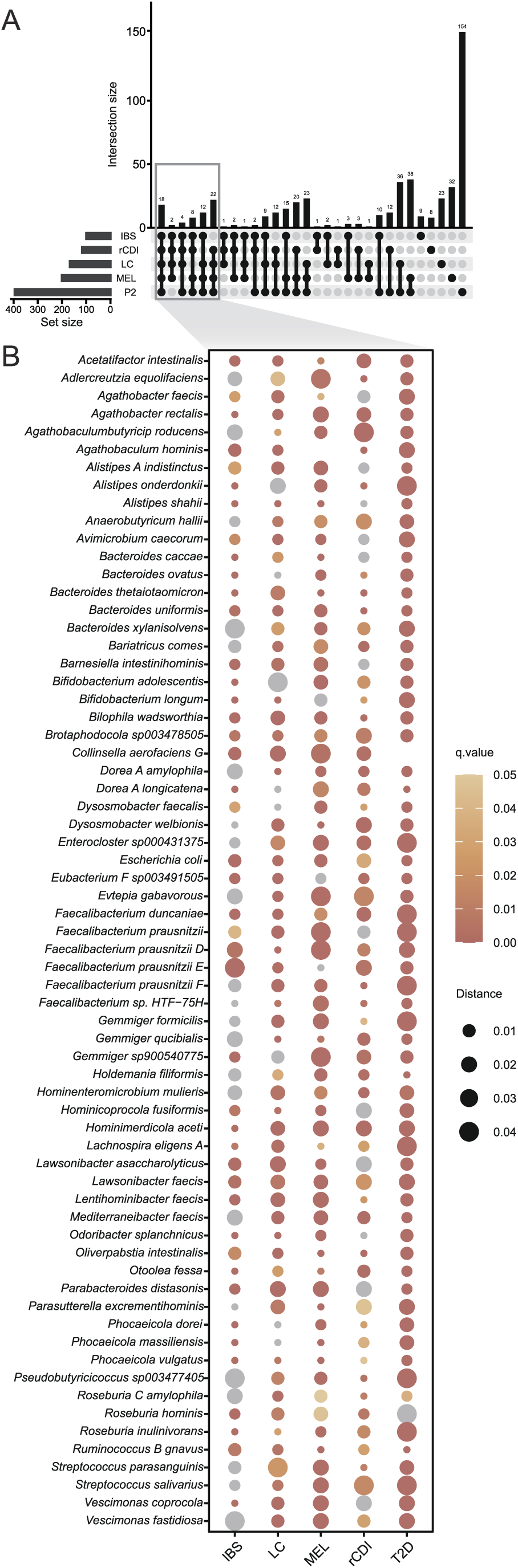
Cross-cohort assessment of population heterogeneity. **A.** UpSet plot showing species with subspecies heterogeneity across 5 datasets. Gray boxes highlight species with significant differences in ≥4 datasets. **B.** Detailed view of 66 species with subspecies heterogeneity in ≥4 datasets. Color scale represents q-values; solid circle with q>0.05 are gray. Solid circle size reflects genetic distance corresponding to q-value.

**Supplementary Figure 12.**
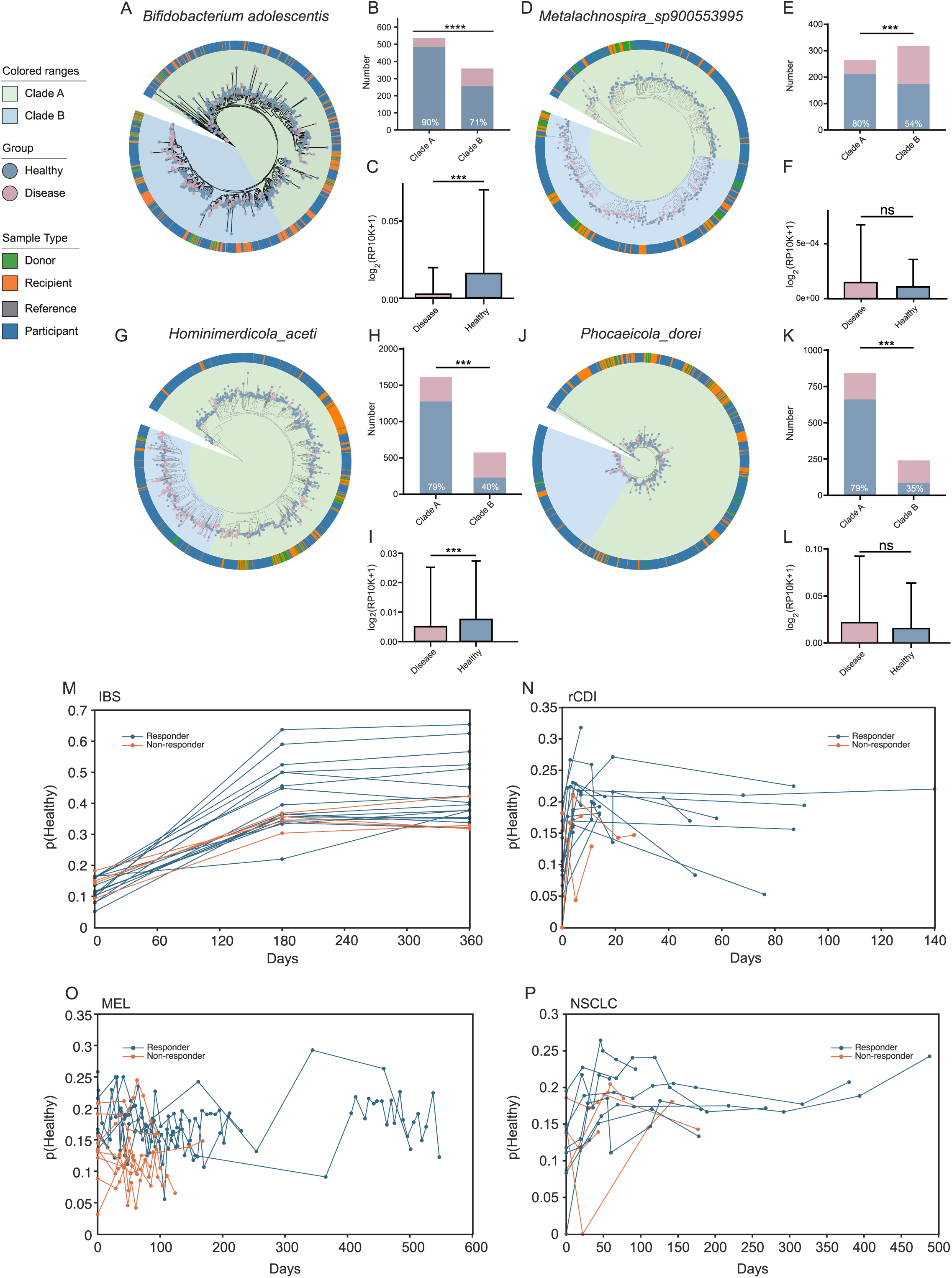
Additional Phylogenetic Analyses and HSIM Scoring. **A–J.** USCG-based phylogenetic analyses of (A-C) *Bifidobacterium adolescentis*,(D-F) *Metalachnospira sp900553995*, (G-I) *Hominimerdicola aceti*, (J-L) *Hominimerdicola_dorei* across 8 cohorts. **M-P.** HSIM scores over time across diseases. (M) IBS, (N) rCDI, (O) MEL, (P) NSCLC.

